# Study of Ancestry, Health, Environment, and Late-Life Neurodegeneration (SAHEL)

**DOI:** 10.64898/2026.09.23.26363703

**Authors:** Suleiman Hamidu Kwairanga, Halima Jafiya Iwar, Amina Umar Faruk, Ibrahim Abdu Wakawa, Umar Baba Musami, Placidus Nwankuba Ogualili, Mohammed Yusuf Mahmood, Muhammad Abba Fugu, Mohammed Mala Gimba, Muktar Mohammed Allamin, Zaharadeen Umar Abbas, Muhammad Kawu Sunkani, Zainab Bukar Yaganami, Fatima Mustapha Kadau, Nasir Muhammad Sani, Peter Danmallam, Luka Emmanuel Nanjul, Larema Babazau, Zaid Muhammad, Dawoud Usman, Abdulrahman Idris Alkhamis, Baba Waru Goni, Babagana Kundi Machina, Cecilia S. Lee, Ibrahim Alkali Allamin, Adam Mustapha, Murtala Bindawa Isah, Salihu Joji, Olugbenga Oguntunde, Yaw Aniweh, Robert Lee Grossman, Takeshi Yoshimatsu, Udunna Anazodo, Chinedu Udeh-Momoh, Thomas K. Karikari, Celeste M. Karch, Chiadi U. Onyike, Mahmoud Bukar Maina

## Abstract

**INTRODUCTION:** Dementia research in sub-Saharan Africa remains limited despite rapid population aging and substantial ancestral, environmental, and vascular diversity. The Study of Ancestry, Health, Environment, and Late-Life Neurodegeneration (SAHEL) characterizes late-life cognitive decline and neurodegeneration in north-eastern Nigeria.

**METHODS:** SAHEL comprises community and hospital cohorts of adults aged 55 years or older in Yobe and Borno States. Participants undergo cognitive, functional, environmental, cardiovascular, metabolic, neuropsychiatric, and neurological assessments. Blood is collected for clinical profiling and biobanking, and consenting hospital participants provide skin biopsies for induced pluripotent stem cell generation. Planned analyses will integrate gut microbiota profiling, retinal imaging, and brain magnetic resonance imaging to investigate biological pathways and markers associated with healthy aging and cognitive outcomes.

**RESULTS:** From August 2025 through February 2026, 1,044 community participants were screened (mean age, 69.4 years; 56.4% female); 379 (36.3%) screened positive for cognitive concern and entered longitudinal follow-up. The hospital cohort enrolled 102 participants (mean age, 73.5 years; 57.8% male), including participants with Alzheimer’s disease, vascular dementia, mixed dementia, and mild cognitive impairment. Blood was collected from 809 community participants and all hospital participants. The cohort includes Kanuri, Fulani, Hausa, Babur/Bura, and Shuwa Arab participants.

**DISCUSSION:** SAHEL has established a large, deeply characterized aging and dementia research platform in north-eastern Nigeria. Its linked clinical, environmental, biomarker, genomic, imaging, microbiome, and cellular resources will enable investigation of population-specific risk and resilience mechanisms and strengthen dementia research and precision medicine in African populations.

## 1. Introduction

Dementia is one of the defining public health challenges of the 21st century. With over 57 million people living with dementia globally and projections exceeding 152 million by 2050, the burden of cognitive decline represents a major medical, social, and economic challenge worldwide [1]. Sub-Saharan Africa is expected to experience one of the most rapid increases in dementia burden globally: by 2050, Africa is projected to account for the largest absolute increase in dementia cases of any world region, driven by demographic aging, high vascular risk factor burden, and limited healthcare infrastructure [2,3]. Yet, the scientific evidence base underpinning cognitive decline and dementia in Africa remains limited. A vast majority of the neurogenerative disease genomics, biomarker, and clinical research were conducted in European-ancestry populations, creating critical knowledge gaps about disease mechanisms, risk factors, and optimal diagnostic and therapeutic strategies in African populations [4]. For instance, the specific ancestral groups of north-eastern Nigeria and the Lake Chad Basin, including the Kanuri, Fulani, Hausa, Babur/Bura, and Shuwa Arab peoples, are virtually absent from the international research literature. These populations have distinct ancestral histories, cultural practices, environmental exposures, and historical migration patterns shaped by longstanding trans-Sahelian migration and admixture processes within the Lake Chad Basin region [5–7], that may influence disease susceptibility, clinical presentation, biomarker profiles, and trajectories of cognitive decline. This dearth of African data perpetuates a cycle of inequity: risk prediction models trained on European ancestry data perform poorly in African populations [8] biomarker thresholds may differ across ancestries, and the environmental and social drivers of cognitive decline and dementia in the African context remain largely uncharacterized [9]. The Lancet Commission on dementia prevention, intervention, and care identified 14 modifiable risk factors collectively accounting for 45% of dementia cases globally [10]. However, the relative contribution of these risk factors within African populations remains poorly understood, particularly in settings where exposure to biomass smoke, contaminated water sources, food insecurity, infectious diseases, and occupational toxicants differ substantially from those in high-income countries [10]. Although several African aging studies have contributed important epidemiological data, including the Indianapolis-Ibadan Dementia Project [11], the Health and Aging in Africa: A longitudinal Study of an INDEPTH Community in South Africa (HAALSI) [12], and the 10/66 Dementia Research Group studies [13], substantial gaps remain in the representation of ancestrally diverse populations from the Lake Chad Basin and north-eastern Nigeria.

The Study of Ancestry, Health, Environment, and Late-Life Neurodegeneration (SAHEL) was designed to address these gaps head-on. Named for an ecological and cultural zone that has nurtured human civilization for millennia, the SAHEL Study integrates epidemiological surveillance, cognitive phenotyping, environmental exposure assessment, cardiovascular and metabolic profiling, and biospecimen collection and analysis within a multidomain dementia research platform. The study aims to elucidate mechanisms linking ancestry and environment to neurodegeneration, identify potentially important risk and resilience factors, and support future mechanistic and biomarker development studies in African populations. This paper describes the rationale, design, recruitment strategy, tools, biospecimen protocols, and baseline characteristics of the SAHEL Study cohort. We present data from both the community-based and hospital-based arms, documenting the scientific quality of the cohort and the exceptional opportunity it represents for global collaborative research. Because most participants in the large community-based arm are not expected to have dementia, the cohort also represents a substantial opportunity for research on healthy and pathological aging and resilience more broadly; and its prospective design will yield incident cases of cognitive decline and dementia that are of particular value for risk-factor research, complementing the prevalent cases identified at baseline.

## 2. Study Design and Conceptual Framework

### 2.1 Overall Design

The SAHEL Study is a prospective, multi-arm cohort study incorporating both cross-sectional baseline assessment and annual longitudinal follow-up. The two arms are complementary: (1) a population-based community cohort enrolling adults aged ≥55 years in Yobe State, north-eastern Nigeria, aimed at characterizing the burden and determinants of cognitive impairment and dementia-related risk factors within the general older population, including environmental, vascular, metabolic, social, and ancestry-related exposures; and (2) a hospital-based clinical cohort recruiting older adults presenting to the Federal Neuropsychiatric Hospital Maiduguri (FNPHM) in Borno State, providing comprehensive clinical phenotyping of neurodegenerative disease cases including characterization of dementia subtypes, neuropsychiatric symptoms, neurological signs, comorbidities, and caregiver burden. Both arms share harmonized data collection instruments, biospecimen protocols, and a common data architecture, enabling integrated analyses across community and clinical settings.

### 2.2 Conceptual Framework

The SAHEL Study is grounded in a life-course, systems-biology model of late-life neurodegeneration, conceptualizing cognitive decline as the cumulative product of decades-long interactions among five intersecting domains. The first is the ancestral and genomic landscape: north-eastern Nigerian populations carry population-specific allele frequencies and epigenetic signatures shaped by founder effects, migration history, and admixture that are almost entirely absent from neurodegenerative disease genetics research. The second is environmental exposure: participants live in an environment characterized by indoor air pollution from biomass combustion, geo-climatic factors like heat stress, recurrent flooding and drought spells, socio-economic and psychological burden of protracted armed conflict, heavy metal and pesticide contamination through water and agricultural produce, and proximity to industrial chemical sources, exposures that are hypothesized to interact with genomic susceptibility across the life course. [14–16] The third encompasses vascular and metabolic trajectories: hypertension, diabetes mellitus, dyslipidemia, and obesity operate as mediators of cerebrovascular and amyloid pathology, with hypertension the leading comorbidity in our hospital setting [17], reflecting a distinctive vascular risk profile of this population. The fourth, social and cultural determinants, encompasses education, occupation, social engagement, religious practice, and traditional lifestyle patterns which operate as modifiers of cognitive reserve and resilience within a cultural context that requires purpose-built measurement approaches. The fifth is neuroinflammatory and infectious pathways: endemic infectious diseases including malaria and helminth infections are hypothesized to prime chronic neuroinflammatory cascades relevant to Alzheimer’s and other neurodegenerative pathologies, a pathway that remains almost entirely unstudied in sub-Saharan African populations. Together, these five domains define a causal framework that cannot be tested in European-ancestry cohorts and SAHEL is positioned to generate findings of direct relevance to African populations worldwide. This conceptual framework is illustrated in Figure 1.

**Figure 1.**
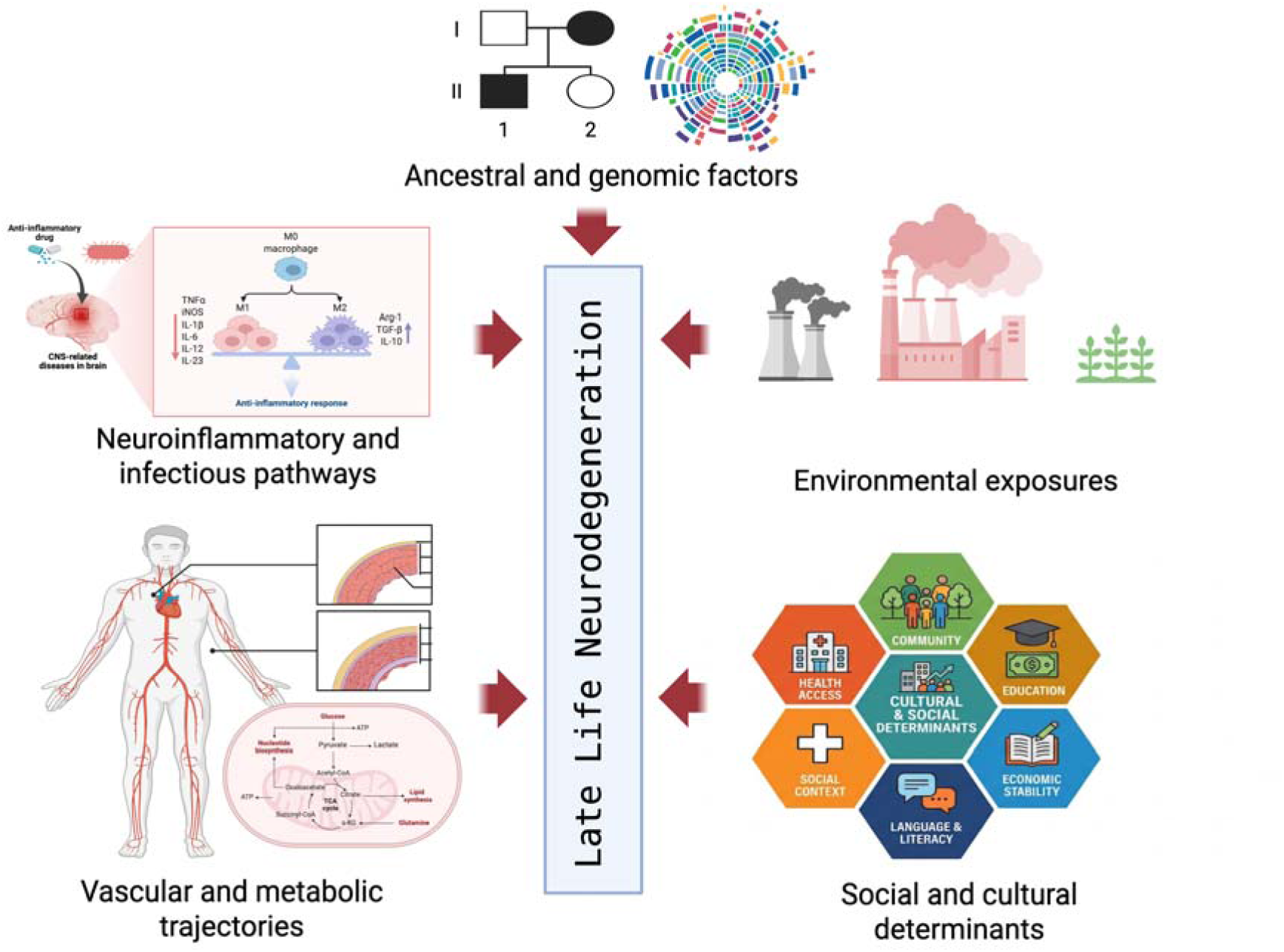
Conceptual framework of the SAHEL Study. Late-life cognitive decline (neurodegeneration) is modeled as the cumulative product of decades-long interactions among five intersecting domains: ancestral and genomic factors, environmental exposures, vascular and metabolic trajectories, social and cultural determinants, and neuroinflammatory and infectious pathways (see Section 2.2).

A related pathway, planned for investigation once longitudinal follow-up specimens become available, is the gut microbiota: environmental neurotoxicant exposure and dietary pattern are established modulators of gut microbial composition, and recent data suggest gut microbiota may moderate the neurodegenerative effect of APOE ε4, a question that has not yet been investigated in any African population[18]

This ancestral diversity also extends to ophthalmic disease genetics: age-related macular degeneration and glaucoma are the leading causes of irreversible blindness globally, their relative prevalence differs markedly between European and African populations, and African-ancestry genetic data for both conditions remain virtually absent from the literature a gap this cohort is positioned to address.

## 3. Study Setting

### 3.1 Yobe State (Community Arm)

The community arm is based in Damaturu, the administrative capital of Yobe State, Nigeria, a state characterized by a semi-arid Sahelian ecology and a diverse ethnic mosaic including Kanuri, Hausa, Fulani, and other indigenous groups. Yobe State shares borders with the Republic of Niger to the north, Borno State to the east, Gombe State to the south, and Bauchi and Jigawa States to the west, reflecting a population shaped by longstanding patterns of migration, trade, and cultural exchange across the Lake Chad Basin region. These multiple borders place Yobe State at a crossroads of ethnocultural exchange, with population movement and intermarriage across all four neighbouring states and Niger contributing to the ethnic and linguistic diversity described above. The region has experienced significant humanitarian challenges in recent years, including displacement, food insecurity, and disruption of health services due to conflict. These challenges highlight the value of local data for describing and contextualizing risk factors for brain aging, cognitive health, and dementia risk in historically underrepresented populations.

### 3.2 Borno State (Hospital Arm)

The hospital arm is anchored at the Federal Neuropsychiatric Hospital Maiduguri (FNPHM), the only premier tertiary neuropsychiatric facility serving Borno State and the broader Lake Chad Basin region. FNPHM is the sole dedicated neuropsychiatric hospital in north-eastern Nigeria, serving a population of approximately 26 million people across multiple states and neighboring countries. [19]. The hospital provides outpatient and inpatient neuropsychiatric services and represents one of the few specialist centers for dementia assessment and management within the region.

## 4. Methods

### 4.1 Community Arm: Eligibility and Recruitment

Participants were eligible for inclusion if they were 55 years or older; resident in the study catchment area (four contiguous wards of Damaturu Local Government Area: Bindigari Pawari, Njiwaji/Gwange, Damaturu Central, and Nayinawa) for at least six months prior to recruitment; willing and able to provide informed consent, or where cognitive capacity was limited, provide assent via proxy obtained from a family member; and residing in a consenting household. Participants were excluded if they had an acute illness or medical emergency that precluded participating at the time of screening; unable to communicate in any of the study languages (Hausa, Kanuri or English); and refused to provide informed consent.

Prior to recruitment, a systematic household listing and mapping exercise was conducted using the QField application software. The GIS team assigned household identification numbers and captured GPS coordinates, while community mobilisers and local leaders conducted sensitization activities to foster community engagement and trust. Pre-consent contact details were stored in a separate database from research data to preserve confidentiality. Assigned household numbers were marked on exterior walls to facilitate identification and follow-up.

Because the study protocol included procedures such as blood sampling and skin biopsy collection that require sterile conditions and specialized handling not readily achievable within participants’ homes, participants were transported to designated Primary Health Care (PHC) facilities rather than assessed within households. Dedicated workstations equipped with biosafety materials were provided at each PHC site, where consent, interviews, cognitive screening, and biospecimen collection were conducted under sterile conditions before immediate sample transport to the laboratory. Transportation support was provided where needed, while community leaders and PHC staff assisted in informing participants about the study and coordinating their attendance at PHC assessment sites. These measures were intended to reduce practical barriers to participation and improve attendance. Data collection was conducted in August 2025, across 999 households, capturing over 1,000 participants with data collected electronically using KoBoCollect deployed on tablet devices.

### 4.2 Community Arm: Instruments and Data Collection

Cognitive screening was performed using the Mini-Cog, comprising three-word recall and the Clock Drawing Test (CDT), generating scores from 0-5; participants scoring 0-2 were classified as screen-positive for cognitive concern and enrolled into the longitudinal cohort, while scores of 3–5 were classified as screen-negative for cognitive concern, consistent with previously reported thresholds used in low-literacy and culturally diverse populations [20,21]. The Mini-Cog was selected because of its brevity, feasibility in low-resource field settings, and prior use in populations with limited formal education. Hausa and Kanuri language versions were developed, tested and refined prior to study start. The Mini-Cog was administered in participants’ preferred languages (English, Hausa, or Kanuri) by field staff fluent in that language. Cognitive domains assessed included memory, visuospatial ability, executive function, and overall Mini-Cog performance. Functional status was assessed using the six-item Basic Activities of Daily Living (BADL) scale, while depressive symptoms were evaluated using the two-item Patient Health Questionnaire (PHQ-2). Standardized cardiovascular and metabolic measurements included blood pressure, height, weight, body mass index (BMI), random blood glucose (RBG), and lipid profile assays from venous blood samples. Environmental exposure assessment captured drinking water source and treatment, cooking fuel and indoor smoke exposure, proximity to environmental hazards, occupational toxicant exposure, dietary exposure pathways, and tobacco use. Detailed sociodemographic and ancestry-related and sociodemographic information including age, sex, ethnicity, educational attainment, occupation, religion, geographic origin and residence, and family history of dementia or memory loss were also systematically collected.

The Mini-Cog was used as a two-stage case-finding instrument rather than a single-stage case identification tool. Participants who screened positive (Mini-Cog score 0–2), together with a randomly sampled subset of those who screened negative (Mini-Cog score 3–5), were selected for subsequent detailed clinical, cognitive and functional assessment. The second-stage assessment was designed to characterize cognitive status, including MCI and dementia where diagnostic criteria were met, and to permit evaluation of the sensitivity, specificity and overall performance of the Mini-Cog in this population. The number of screen-negative participants selected will be determined by the number of participants screening positive, with an approximately 1:1 random sample of screen-negative participants selected for second-stage assessment.

To provide a comprehensive assessment of domain-specific cognitive functioning at the second wave, all participants in the community cohort will undergo a neuropsychological test battery (NTB). The NTB was selected in part because of concerns regarding the suitability of the Mini-Cog for assessing longitudinal cognitive change in this population. Performance on the clock-drawing component may be influenced by literacy and educational experience, potentially resulting in low scores that do not reflect cognitive decline. Preliminary analysis of baseline data supports this concern: 82.8% of participants (864/1,044) scored zero on the clock-drawing component, and clock-drawing failure was strongly associated with educational attainment (89.5% among participants with Quranic-only education vs. 40.0% among those with any formal schooling; χ²=91.1, p<0.001), consistent with this item capturing literacy and numeracy rather than cognitive impairment in this population. The NTB was therefore selected to provide a more culturally appropriate and cross-culturally applicable assessment of cognitive functioning, while also facilitating harmonisation with the Africa-FINGERS study [22], of which the community cohort site is a member.

The NTB comprises the Animal Naming Test, which assesses verbal fluency and executive and language functions; Word List Recall Trials, which assess verbal learning and memory; the Nigerian Matchstick Test, which assesses visuospatial functioning; Brave Man Story Recall, which assesses episodic memory [23] ; the Multilingual Naming Test, which assesses object-naming ability and language; the Frontal Assessment Battery (FAB), which evaluates executive dysfunction; and the Digit Span Test, which assesses verbal short-term and working memory. The tests will be administered in the following order: Animal Naming trial, first Brave Man Story recall, Word List immediate recall trials, Nigerian Matchstick Test, Word List recall trial, Word List recognition trial, FAB, Digit Span, Brave Man delayed recall, Nigerian Matchstick delayed recall, and Nigerian Matchstick delayed recognition.

The Identification and Intervention for Dementia in Elderly Africans (IDEA) score [24] will be derived from the NTB and used as the screening measure for possible dementia at the second wave. Participants identified as being at risk based on their IDEA score will subsequently undergo the full structured clinical assessment (SCA; section 4.4) administered by a study physician. Thus, the NTB provides detailed, domain-specific measures of cognitive functioning, while the IDEA score provides a standardized approach for identifying participants who may require further clinical assessment. The cognitive measures were selected for their cross-cultural utility and were translated to Hausa language.

The Mini-Cog administered at baseline will also provide an opportunity to compare the performance of the baseline screening measure with the more detailed cognitive assessment conducted at the second wave. This comparison will allow us to examine the relationship between the Mini-Cog and the NTB/IDEA assessment and to assess whether the former has any utility for brief population screening for cognitive dysfunction in this population.

### 4.3 Hospital Arm: Eligibility and Recruitment

Participants were eligible for inclusion if they were similarly 55 years or older; presenting to or admitted at FNPHM Maiduguri due to a complaint of or referral for cognitive decline, memory impairment or suspected dementia; availability of an informant (family member or caregiver) for corroborative history and proxy-related instruments.

Hospital participants were recruited consecutively from outpatient neurology and psychiatry clinics and inpatient wards at FNPHM, between September 2025 and February 2026. Recruitment was conducted by specialist clinicians including neurologists and neuropsychiatrists with expertise in dementia assessment. Recruitment is still ongoing.

### 4.4 Hospital Arm: Instruments and Data Collection

Hospital arm participants underwent, in addition to clinical interview and neurological examination, a comprehensive, multi-domain Structured Clinical Assessment (SCA) where cognitive function was evaluated using established psychometric assessments. The Montreal Cognitive Assessment (MoCA; score range 0–30), provides a summary assessment of cognitive status covering visuospatial/executive function, naming, memory, attention, language, abstraction, and orientation, with visuospatial tasks photographically documented for quality assurance review.[25,26] Dementia severity was rated by caregivers using the Quick Dementia Rating System (QDRS; 0–30), [27] and neuropsychiatric symptoms were captured using a 14-item Modified Neuropsychiatric Inventory Questionnaire (NPI-Q) (expanded by the study team from the standard 12-item version to include two additional locally relevant symptom items), scored for both presence and severity. [28] Functional independence was assessed with the Lawton-Brody Instrumental Activities of Daily Living scale (IADL; 0-8), and caregiver burden with the 12-item Zarit Burden Interview (ZBI-12; 0-48).[29,30] All consenting participants underwent a structured bedside neurological examination by the specialists, assessing consciousness, cranial nerves, motor system, tremor, sensory modalities, gait, and speech, with findings digitally recorded and GPS geo-tagged. Primary and secondary diagnoses were assigned by specialist clinicians using ICD-10 criteria, spanning Alzheimer’s disease, vascular dementia, mixed dementia, Lewy body dementia, frontotemporal dementia, and mild cognitive impairment, with clinician diagnostic confidence rated on a 0–10 scale. Comorbidities were also systematically recorded.

The NTB will also be administered to participants in the hospital arm. However, unlike participants in the community arm, all participants in the hospital arm will undergo the SCA.

### 4.5 Biospecimen Collection and Processing

#### 4.5.1 Community Arm

Venous blood was collected from all consenting community participants by trained phlebotomists using standardized protocols. Three vacutainer types were employed: EDTA vacutainers for whole blood preservation, enabling complete blood count analysis and buffy coat archiving for future genomic DNA extraction; plain vacutainers for serum separation, supporting lipid profile quantification, serological testing, and biomarker assays; and fluoride oxalate vacutainers for plasma glucose measurement. Out of the 1,044 community participants, 809 (77.5%) had their venous blood collected. Blood samples were not obtained from 235 (32.5%) participants due to either difficulty in getting venous samples or refusal to consent for phlebotomy. Blood samples were processed in the field within two hours of collection using portable centrifuges operating at 3,000 rpm for ten minutes. Separated serum and plasma aliquots were stored in cryovials at -20°C in field-deployed ultra-cold storage units before transfer to the central biobank within 24 to 48 hours of collection. Sample transport and storage temperatures were monitored continuously throughout field operations to minimise pre-analytical variability. Buffy coat layers were archived separately for future genomic DNA extraction. All specimens were labeled with unique participant identifiers to enable precise chain-of-custody tracking throughout the biobanking process. Analytes measured from community biospecimens included lipid profile (total cholesterol, LDL cholesterol, HDL cholesterol, and triglycerides), random blood glucose, and serological serum markers for common chronic viral infections (i.e. HIV, HBV and HCV); which results were reported separately. Cardiovascular risk was flagged using a composite assessment of lipid profile, glucose, and blood pressure.

#### 4.5.2 Hospital Arm

All 102 hospital participants underwent venous blood sampling using the same vacutainer protocol as the community arm. Samples were processed immediately at the Federal Neuropsychiatric Hospital Maiduguri (FNPHM) laboratory, with all analytes assayed within four hours of collection. Residual serum and plasma aliquots were cryopreserved at minus 20 degrees Celsius in the hospital biobank for future analyses. Buffy coat was archived from all hospital participants for future genomic DNA extraction.

#### 4.5.3 Skin Biopsies and iPSC Generation

Skin biopsies were collected from consenting community and hospital participants by trained clinicians under sterile conditions. Each 3 mm punch biopsy was placed immediately into fibroblast growth medium and transported within five hours to the Biomedical Science Research and Training Centre (BioRTC), a state-of-the-art research facility established at Yobe State University in partnership with the Yobe State Government. At BioRTC, fibroblasts are cultured, expanded, and reprogrammed into induced pluripotent stem cells (iPSCs) using established episomal reprogramming protocols [31]

Generated iPSC lines will be deposited in the African Somatic and Stem Cell Bank, an open-access biobank distributed across BioRTC (Nigeria), the University of Sussex (UK), and Washington University in St. Louis (USA) [43]. This distributed biobanking architecture provides redundancy, international accessibility and long-term preservation of a resource that, to our knowledge, represents the first open-access iPSC biobank established specifically for dementia research in an African population. iPSC lines will be differentiated into disease-relevant brain cell types, including neurons, astrocytes and microglia, and into cerebral organoids, to investigate how genetic and ancestral variation influences cellular and molecular pathways relevant to Alzheimer’s disease and related dementias. Initial studies will examine APOE-associated biology, while the resource will enable investigation of additional genetic risk and protective factors as they emerge.

#### 4.5.4 Biobank Quality Assurance

All field and laboratory biospecimen handling adhered to ISO 20387-compliant procedures for biological resource centers. Quality assurance measures included pre-analytical temperature monitoring of sample tubes from collection through centrifugation, duplicate aliquoting to minimize freeze-thaw degradation, and sample tracking at each transfer point. Biobank storage temperatures were audited regularly, and formal standard operating procedures were documented for all collection, processing, aliquoting, and storage steps.

#### 4.5.5 Stool Collection and Gut Microbiome

Stool specimens will be collected using sterile self-collection kits and stored at −80°C pending microbiome analysis. Bacterial community composition will be characterized using 16S rRNA gene sequencing to assess microbial diversity and taxonomic profiles. These data will enable investigation of relationships between the gut microbiota, dietary and environmental exposures, genetic background, and cognitive and clinical phenotypes within the cohort, providing a basis for future investigation of the gut–brain axis in this underrepresented population.

#### 4.5.6 Ophthalmic and Retinal Phenotyping

Comprehensive ophthalmic assessment will include retinal imaging using the Topcon Maestro2 platform (Topcon Healthcare, Japan), incorporating three-dimensional macular (6×6mm) and wide-field (12×9mm) optical coherence tomography (OCT) imaging, OCT angiography (3×3mm), and color fundus photography. These modalities will enable quantitative characterization of retinal structure and microvasculature, alongside assessment of prevalent ocular pathology. Retinal phenotypes will be integrated with cognitive, clinical, biomarker, and genetic data to investigate relationships between ocular and retinal changes, systemic and neurodegenerative disease, and genetic variation within the cohort.

#### 4.5.7 Magnetic Resonance Imaging (MRI)

The enrolled participants will undergo brain imaging using a 1.5-T magnetic resonance imaging (MRI) scanner at the University of Maiduguri Teaching Hospital. Image acquisition will follow the Africa Dementia Imaging Protocol (ADIP), adapted from established Alzheimer’s Disease Neuroimaging Initiative protocols for use in Africa-FINGERS. [22] ADIP provides a comprehensive assessment of regional brain volume, cortical thickness, cerebral blood flow, white-matter hyperintensity burden, and markers of structural and functional connectivity. These measures will be used to investigate associations among neurodegeneration, dementia severity, and cerebrovascular disease in the SAHEL hospital cohort.

### 4.6 Data Management and Quality Assurance

All data were collected using KoBoCollect deployed on encrypted tablet devices, enabling real-time data entry, automated range checks, and mandatory field completion for clinically critical variables. GPS coordinates were recorded at the household or interview-site level to support spatial analyses of environmental exposure. Data were transferred via encrypted server connection to a secure, password-protected database hosted on institutionally approved infrastructure. All personally identifiable information was stored separately from study data and linked only through unique participant identifier codes to ensure participants’ confidentiality. Data quality was monitored continuously by a central data management team through automated weekly reports covering missing data rates, out-of-range values, and interviewer-level consistency checks.

### 4.7 Ethical Considerations

The SAHEL Study received ethical approval from the Federal Neuropsychiatric Hospital Maiduguri’s Institutional Research and Ethical Board (Reference: FNPH/082023/REC140) and the Yobe State Ministry of Health’s State Health Research and Ethical Committee (Reference: MOH/GEN/747/Vol. 1). All participants provided written informed consent prior to enrolment; for participants who were not literate, consent was obtained via thumbprint impression, witnessed by an independent literate witness, following standard Good Clinical Practice guidance; where participants lacked capacity to consent, written proxy consent was obtained from the primary caregiver or next-of-kin, and participant assent was sought wherever feasible.

The study was conducted in accordance with the Declaration of Helsinki, the Nigerian National Code for Health Research Ethics, and Good Clinical Practice guidelines. Community and religious leaders were engaged prior to study commencement to build trust and facilitate culturally sensitive recruitment. All field interviews and cognitive assessments were conducted in participants’ preferred languages (English, Hausa or Kanuri), with culturally validated adaptations of instruments. Participant remuneration was provided in accordance with local ethical standards to compensate for time without inducing undue influence.

## 5. Baseline Cohort Characteristics

### 5.1 Community Arm

#### 5.1.1 Participation and Enrolment

The SAHEL community arm screened 1,044 older adults across 999 households in four wards of Damaturu LGA, Yobe State (Table 1). Among these, 379 participants (36.3%) screened positive for cognitive concern (Mini-Cog score 0 to 2) and 665 (63.7%) screened negative (Mini-Cog score 3 to 5). All 1,044 participants, regardless of Mini-Cog result, will complete the full NTB and if eligible, a clinical examination that includes the SCA will be performed. These participants will be longitudinally followed, enabling prospective tracking of cognitive trajectories, incident dementia, and the environmental, vascular, and genomic factors that predict conversion from intact cognition to cognitive impairment over time. Participants meeting diagnostic criteria for dementia will be compared against cognitively intact controls drawn from the same community sample. The achieved sample represents a high-density enumeration of the older adult population in these wards, with a household-level coverage reflecting exceptional community engagement and field team performance.

**Table 1.**
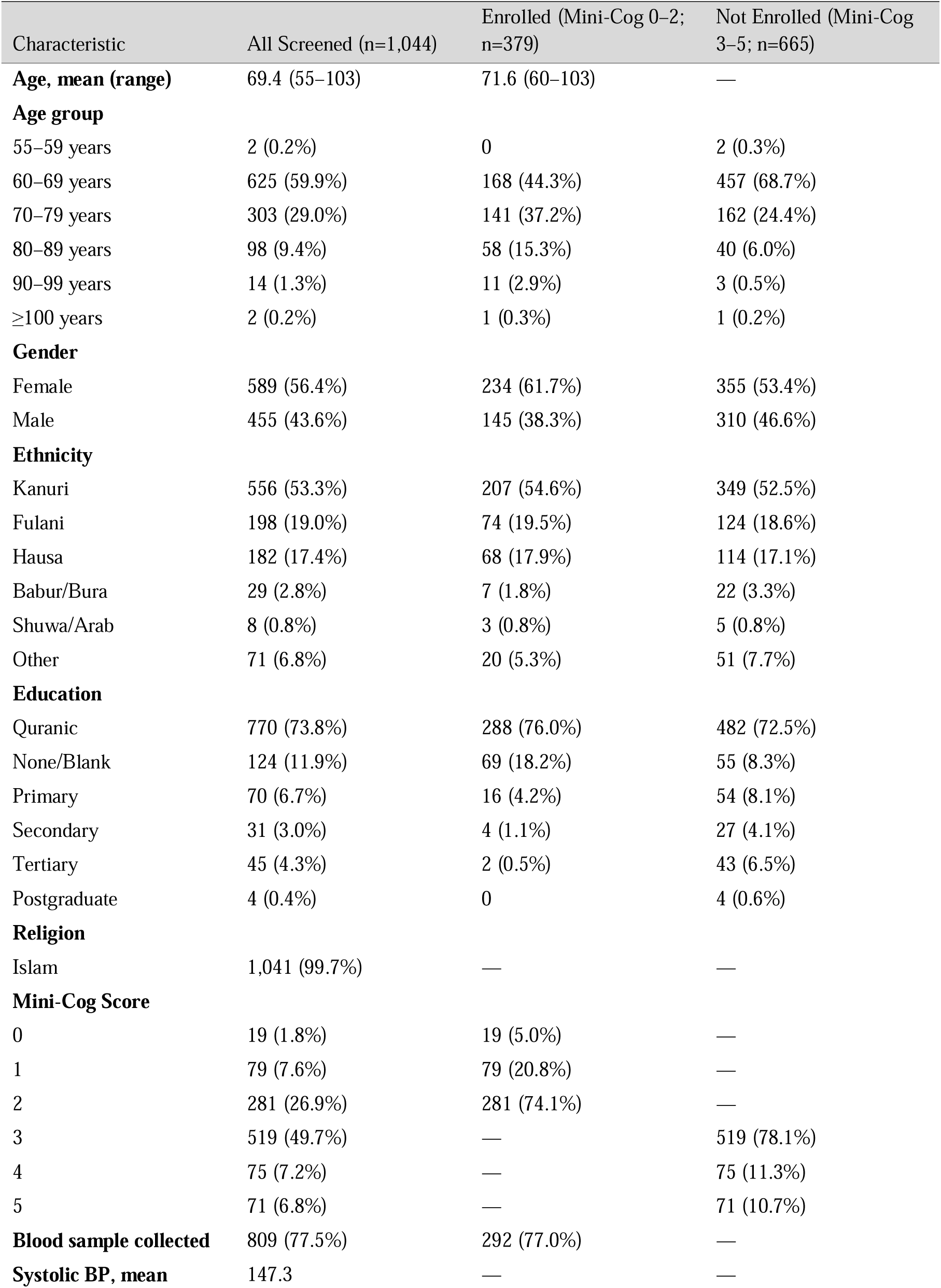
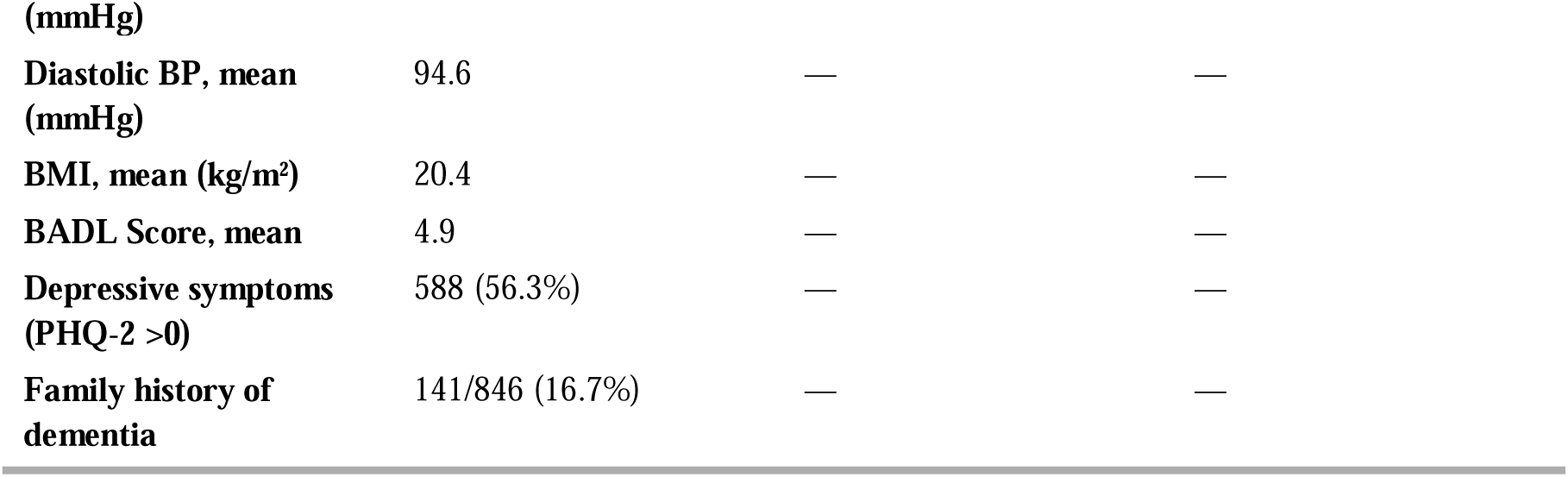
Baseline Characteristics of the SAHEL Community Cohort (n=1,044)

#### 5.1.2 Demographic Characteristics

The community sample had a mean age of 69.4 years (range 55–103), was predominantly female (56.4%), and fell largely within the 60–69 age group (59.9%), with two centenarians identified. Among screen-positive enrolled participants, mean age was 71.6 years with a marked female predominance (61.7%), consistent with the greater longevity and dementia risk of women globally [32,33]. Kanuri was the largest ethnic group (53.3%), followed by Fulani (19.0%), Hausa (17.4%), and smaller minorities including Babur/Bura and Shuwa Arab, a composition uniquely positioned to power ancestry-stratified genetic analyses. Educational attainment was predominantly Quranic (73.8%), with only 10.9% having attended formal secular schooling, a pattern with important implications for cognitive reserve modeling. The predominant occupations were trading (41.4%), farming (22.9%), and unemployed/retired (17.5%), reflecting the agrarian and informal-economy livelihoods of north-eastern Nigeria. (Figure 2A–B).

**Figure 2.**
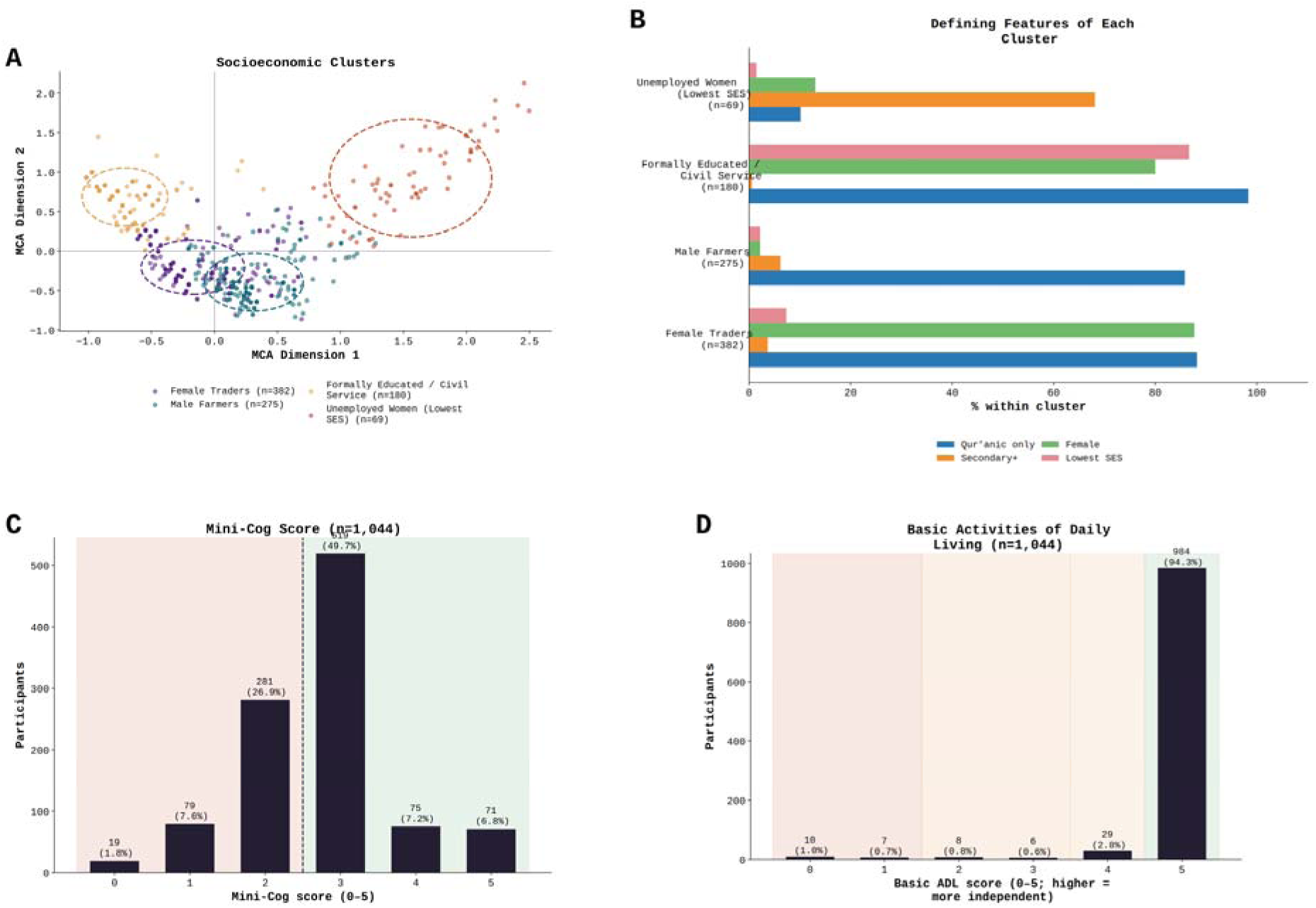
Socioeconomic clusters, cognitive screening, and functional status in the SAHEL community cohort. (A) Multiple correspondence analysis (MCA) showing the distribution of participants across four socioeconomic clusters: female traders (n = 382; 42%), male farmers (n = 275; 30%), formally educated/civil-service workers (n = 180; 20%), and unemployed women with the lowest socioeconomic status (SES; n = 69; 8%). Dashed ellipses indicate the concentration of each cluster in MCA space. (B) Percentage of participants within each cluster characterized by Qur’anic-only education, secondary or higher education, female sex, and the lowest SES (class V). (C) Mini-Cog score distribution; scores of 0–2 were classified as screen-positive and scores of 3–5 as screen-negative. Thirty-six percent of screened participants (n = 379) were enrolled in the longitudinal cohort based on a Mini-Cog score of 2 or lower. (D) Basic activities of daily living (BADL) score distribution; the median BADL score was 5, and 94% of participants were fully independent.

**Figure 3.**
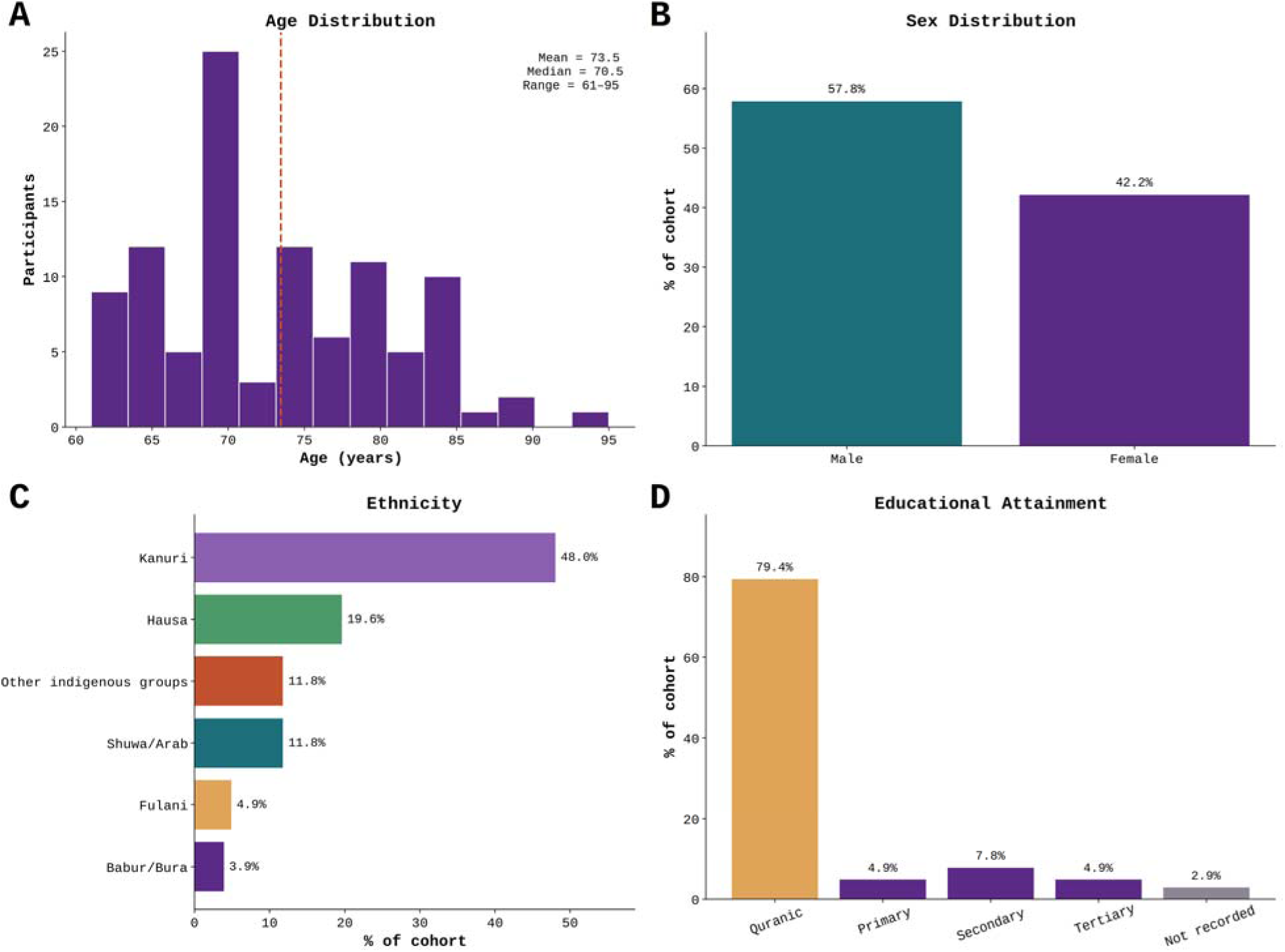
Demographic characteristics of the SAHEL hospital cohort. (A) Age distribution of the 102 participants enrolled at NNDR Hospital; the mean age was 73.5 years, the median was 70.5 years, and the range was 61–95 years. The dashed line indicates the mean. (B) Sex distribution: 59 participants (57.8%) were male and 43 (42.2%) were female. (C) Ethnic composition; Kanuri participants constituted the largest group (48.0%), followed by Hausa participants (19.6%). (D) Educational attainment; Qur’anic education was the predominant category (79.4%).

#### 5.1.3 Cognitive Screening Results

The Mini-Cog score distribution across the full community sample was: score 0 (n=19, 1.8%), score 1 (n=79, 7.6%), score 2 (n=281, 26.9%), score 3 (n=519, 49.7%), score 4 (n=75, 7.2%), and score 5 (n=71, 6.8%). Scores 0-2 were the criterion for enrolment (positive screen), yielding an overall cognitive concern prevalence of 36.3% in this older population. Together with a randomly sampled subset of screen-negative participants, screen-positive participants were selected for further clinical cognitive assessment, as described in Section 4.2. This high prevalence likely reflects the high-risk sociodemographic profile of the screened population, including limited formal education, high vascular risk factor burden, and environmental exposures. The proportion of participants that screened positive for cognitive concern is comparable to findings reported in several previous African aging studies using community-based cognitive screening approaches [34–36].

Mean Basic Activities of Daily Living (BADL) score in the community was 4.9 out of 5.0, indicating that most screened participants remained functionally independent in basic self-care, providing an important baseline against which future functional decline can be tracked.

#### 5.1.4 Cardiovascular and Metabolic Profile

The community cohort carries a substantial cardiovascular risk burden. Mean systolic blood pressure was 147.3 mmHg and mean diastolic blood pressure was 94.6 mmHg, indicating a substantial burden of elevated blood pressure and reflecting the broader epidemiological transition toward non-communicable diseases, including hypertension, documented across sub-Saharan Africa [44]. Mean BMI was 20.4 kg/m², indicating a predominantly lean or normal-weight population, reflecting the nutritional context of north-eastern Nigeria. Serum lipid profile, random blood glucose, and serology results for common viral infections in the biomarker subsample are summarized in Figures 5–7.

**Figure 4.**
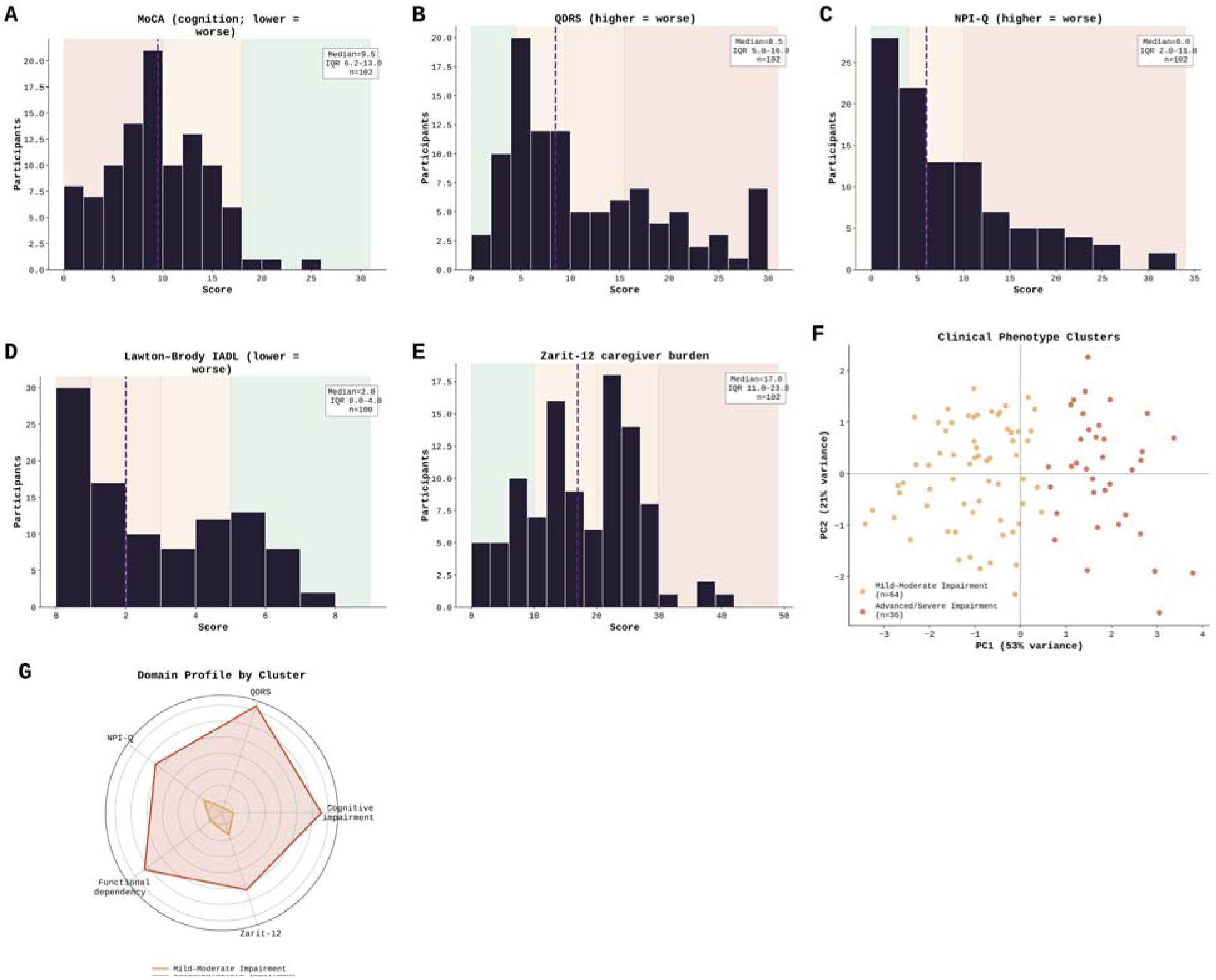
Clinical-domain distributions and phenotype clusters in the SAHEL hospital cohort. (A) Montreal Cognitive Assessment (MoCA) scores. (B) Quick Dementia Rating System (QDRS) scores. (C) Neuropsychiatric Inventory Questionnaire (NPI-Q) scores. (D) Lawton– Brody instrumental activities of daily living (IADL) scores. (E) Twelve-item Zarit Burden Interview (Zarit-12) scores. In panels A–E, dashed lines indicate median values, annotation boxes report medians and interquartile ranges, and shaded regions represent established severity categories. Lower MoCA and Lawton–Brody IADL scores indicate greater impairment or dependency; higher QDRS, NPI-Q, and Zarit-12 scores indicate greater disease severity or burden. (F) Principal component analysis of five standardized domain scores among participants with complete data (n = 100), showing mild-to-moderate impairment (n = 64; 64%) and advanced/severe impairment (n = 36; 36%) clusters. The first two principal components explained 53% and 21% of the variance, respectively. (G) Mean standardized domain profiles of the two clusters; greater outward displacement indicates greater impairment, dependency, or burden.

**Figure 5.**
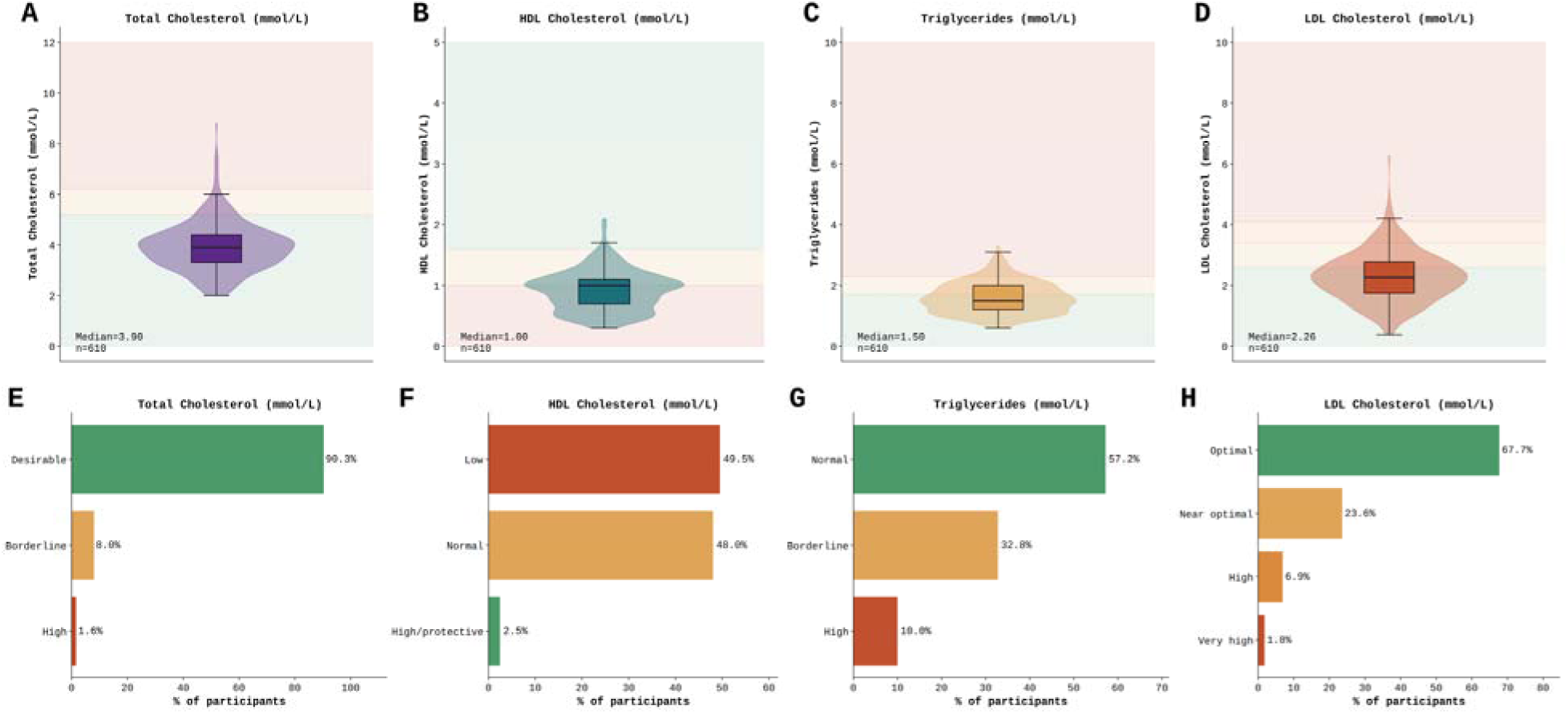
Lipid distributions and clinical classifications in the SAHEL community cohort (n = 610). Violin plots show (A) total cholesterol (TC), (B) high-density lipoprotein (HDL) cholesterol, (C) triglycerides (TG), and (D) low-density lipoprotein (LDL) cholesterol. Horizontal reference lines denote clinically relevant thresholds. Clinical categories are shown for (E) TC, (F) HDL cholesterol, (G) TG, and (H) LDL cholesterol calculated using the Friedewald equation. Overall, 90.3% had desirable TC (<5.2 mmol/L), 49.5% had low HDL (<1.0 mmol/L), 42.8% had borderline-to-high TG, and 67.7% had optimal LDL (<2.6 mmol/L).

**Figure 6.**
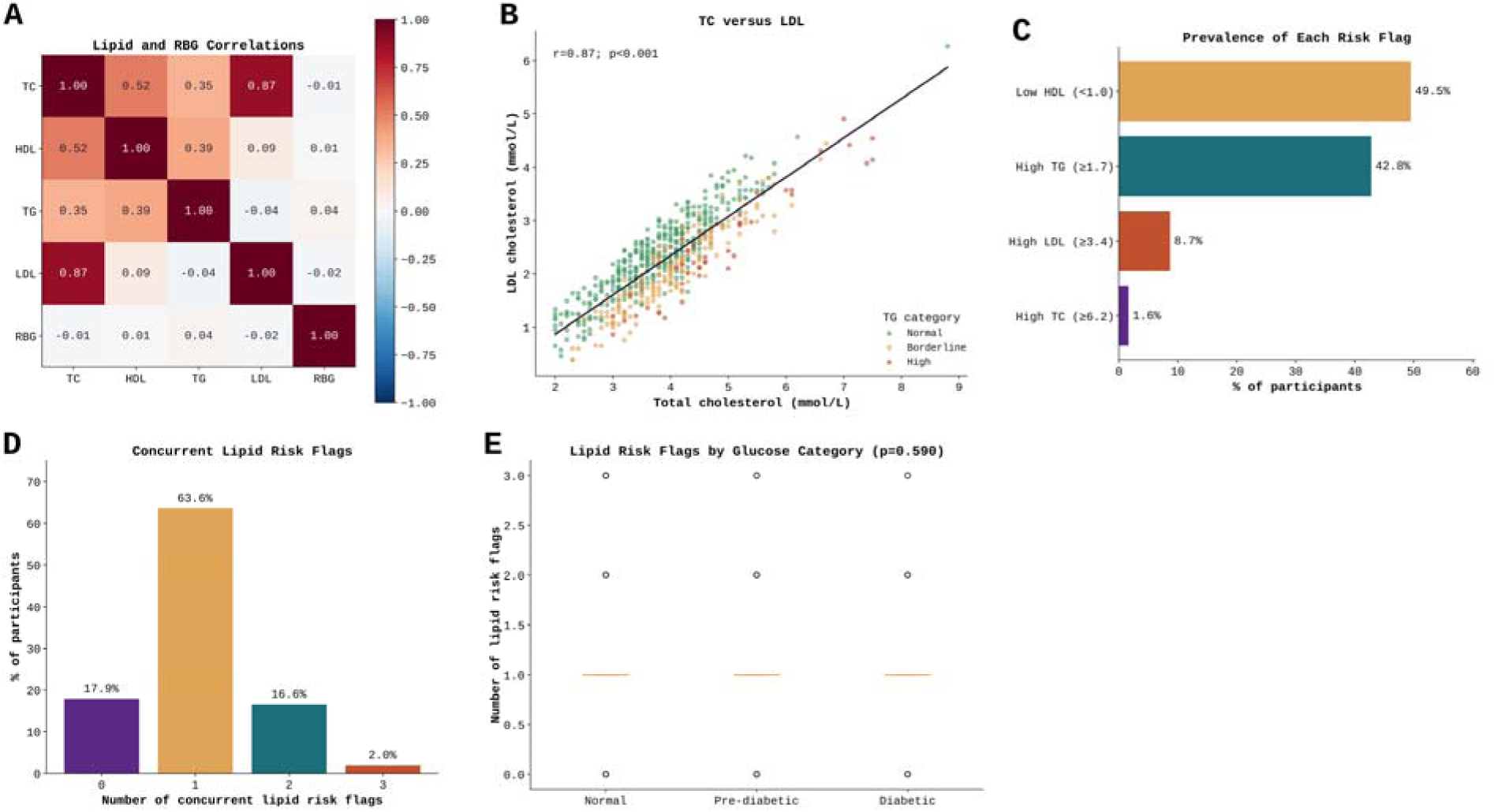
Inter-biomarker correlations and cardiovascular lipid risk-flag burden in the SAHE community cohort. (A) Pearson correlation heatmap for TC, HDL cholesterol, TG, LDL cholesterol, and random blood glucose (RBG). (B) Scatterplot of TC versus LDL cholesterol, color-coded by TG category. (C) Prevalence of individual lipid risk flags: high TC (≥6.2 mmol/L), high LDL (≥3.4 mmol/L), low HDL (<1.0 mmol/L), and high TG (≥1.7 mmol/L). (D) Distribution of concurrent lipid risk flags. (E) Lipid risk-flag counts by glucose category (Kruskal–Wallis p = 0.962).

**Figure 7.**
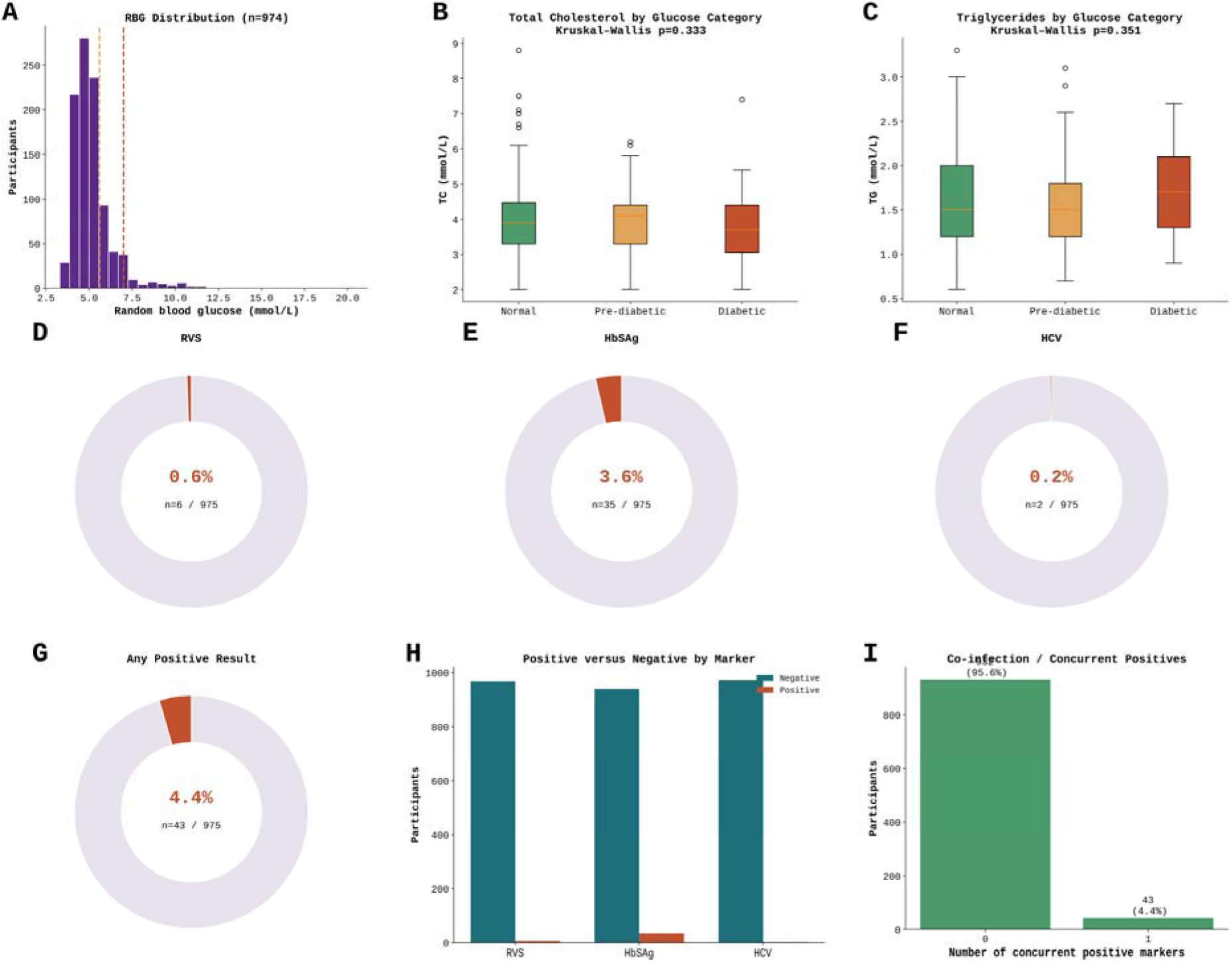
Random blood glucose, lipid associations, and serologic screening in the SAHEL community cohort. (A) RBG frequency distribution. (B) Total cholesterol and (C) triglyceride distributions by glycemic category. Donut charts show the prevalence of (D) RVS positivity, (E) hepatitis B surface antigen positivity, (F) hepatitis C antibody positivity, and (G) any positive serologic result. (H) Positive and negative results by marker. (I) Concurrent positive markers; 95.6% had no positive marker and 4.4% had exactly one, with no observed coinfections.

#### 5.1.5 Environmental Exposures

The community cohort had a substantial prevalence of factors associated with potential environmental exposures (Table 3). The majority of participants (98.7%) relied on groundwater from boreholes as their primary source of drinking water, which may represent a potential route of exposure to contaminants such as heavy metals, fluoride and nitrate [37,38]. Most participants used firewood (91.4%) or charcoal (8.4%) as their primary cooking fuel, both of which generate household air pollutants, including PM2.5, carbon monoxide and polycyclic aromatic hydrocarbons, that have been associated with neuroinflammation and dementia risk [39]. Additionally, 41.1% lived in proximity to potential environmental hazards, such as dumpsites or agrochemical farms, 11.5% reported household or occupational toxicant exposure, and 69.3% regularly consumed locally grown produce. Where produce is irrigated with contaminated water or grown near industrial or waste-disposal sites, consumption may represent a route of exposure to bioaccumulated contaminants [40]. Collectively, these findings identify several potential routes of chronic environmental exposure that warrant direct characterization and investigation in relation to cognitive aging and dementia.

Tobacco use was rare, with 98.9% of participants reporting that they had never smoked. Family history of dementia was reported in 16.7% of participants with available data, providing an opportunity for future investigation of familial aggregation. Depressive symptoms (PHQ-2 >0) were present in 56.3% of participants, indicating a high prevalence of depressive symptoms in the cohort and providing an important context for investigating the established relationship between depression and dementia risk.

### 5.2 Hospital Clinical Arm

Several assessments within the hospital cohort remain ongoing, and some clinical variables were unavailable for all participants at the time of analysis. The SAHEL Clinical arm enrolled 102 participants between September 2025 and February 2026 at FNPHM, with a mean age of 73.5 years (57.8% male, 95.1% Muslim, 79.4% had Quranic education, and 83.3% were in lower occupational social classes, based on the participant’s own occupation rather than a composite household socioeconomic index). Participants were drawn from diverse ethnic groups in the region, with Kanuri being the largest group (48.0%). Clinical assessment revealed severe cognitive impairment (mean MoCA 9.6, range 0–25; mean QDRS 11.4), marked functional dependence (mean Lawton-Brody IADL 2.4 out of 8), high neuropsychiatric symptom burden (mean NPI-Q 7.8), and moderate to high caregiver burden (mean ZBI-12 17.0).

Specialist clinical assessment was completed in 21 of 102 enrolled participants (20.6%) at the time of analysis. Among these, Alzheimer’s disease was the predominant diagnosis (66.7%), followed by vascular dementia (14.3%), mixed dementia (9.5%), and mild cognitive impairment (9.5%). Mean clinician diagnostic confidence was 5.8 out of 10, reflecting moderate confidence in the absence of neuroimaging or cerebrospinal fluid biomarkers. Comorbidities included hypertension (9.8%) and diabetes mellitus (2.0%). Formal neurological examination was completed in 49.0% of participants, with ongoing assessments scheduled for the remainder. Detailed neurological examination findings will be reported separately (Table 2; Figures 2–3).

**Table 2.** Baseline Characteristics of the SAHEL Hospital Clinical Cohort (n=102)

| Characteristic | n (%) or Mean (Range) |
| --- | --- |
| **Age, mean (range)** | 73.5 years (61–95) |
| **Age group** |  |
| 60–69 years | 27 (26.5%) |
| 70–79 years | 46 (45.1%) |
| ≥80 years | 29 (28.4%) |
| **Gender** |  |
| Male | 59 (57.8%) |
| Female | 43 (42.2%) |
| **State of origin** |  |
| Borno State | 72 (70.6%) |
| Yobe State | 10 (9.8%) |
| Kano State | 8 (7.8%) |
| Other states | 12 (11.8%) |
| **Ethnicity** |  |
| Kanuri | 49 (48.0%) |
| Hausa | 20 (19.6%) |
| Shuwa/Arab | 12 (11.8%) |
| Other | 12 (11.8%) |
| Fulani | 5 (4.9%) |
| Babur/Bura | 4 (3.9%) |
| **Education** |  |
| Quranic | 81 (79.4%) |
| Secondary | 8 (7.8%) |
| Primary | 5 (4.9%) |
| Tertiary | 5 (4.9%) |
| None | 3 (2.9%) |
| **Religion** |  |
| Islam | 97 (95.1%) |
| Christianity | 5 (4.9%) |
| **MoCA Score, mean (range)** | 9.6 (0–25) |
| **QDRS Score, mean (range)** | 11.4 (1–30) |
| **QDRS Behavioral Subtotal, mean** | 6.6 |
| **Lawton-Brody IADL, mean (range)** | 2.4 (0–7) |
| **NPI-Q Total Score, mean** | 7.8 |
| **Zarit Burden (ZBI-12), mean** | 17.0 |
| **Blood sample collected** | 102 (100%) |
| **Primary diagnosis (n=21 with complete assessment)** |  |
| Alzheimer's disease | 14 (66.7%) |
| Vascular dementia | 3 (14.3%) |
| Mixed AD + VaD | 2 (9.5%) |
| Mild cognitive impairment | 2 (9.5%) |
| **Comorbidities** |  |
| Hypertension | 10 (9.8%) |
| Diabetes mellitus | 2 (2.0%) |
| Cerebrovascular disease | 0 |
| **Clinical exam completed** | 50 (49.0%) |

**Table 3.** Environmental Exposure Profile, Community Cohort (n=1,044)

| Exposure | n (%) |
| --- | --- |
| **Primary drinking water source** |  |
| Borehole | 1,029 (98.7%) |
| Tap water | 10 (1.0%) |
| Sachet/bottled water | 3 (0.3%) |
| Hand pump / other | 2 (0.2%) |
| **Primary cooking fuel** |  |
| Firewood | 953 (91.4%) |
| Charcoal | 88 (8.4%) |
| Gas | 2 (0.2%) |
| Electricity | 1 (0.1%) |
| **Proximity to environmental hazard** |  |
| Yes (dumpsite / road / industry / agrochemical farm) | 429 (41.1%) |
| No | 554 (53.1%) |
| Not sure | 37 (3.5%) |
| **Occupational toxicant exposure (household)** |  |
| Yes (welding / mining / chemicals) | 120 (11.5%) |
| No | 725 (69.4%) |
| Not sure | 41 (3.9%) |
| **Locally grown vegetable consumption** |  |
| Yes | 723 (69.3%) |
| No | 74 (7.1%) |
| **Tobacco use** |  |
| Never | 1,031 (98.9%) |
| Past smoker | 10 (1.0%) |
| Current smoker | 3 (0.3%) |

## 6. Data Availability, Linkage, and Future Analyses

### 6.1 Data Architecture

All SAHEL data are held in a secure, access-controlled longitudinal database. Participant identifiers are stored separately from analytical datasets and linked only through unique study codes, ensuring both confidentiality and data security. The database is structured into harmonised layers covering: core demographic and sociodemographic variables; cognitive assessment scores from the Mini-Cog, MoCA, QDRS and PHQ-2; measures of function, behavior and caregiver burden, including the Basic Activities of Daily Living scale (BADL), Lawton-Brody Instrumental Activities of Daily Living Scale (IADL), NPI-Q and ZBI-12; environmental exposure data covering water sources, cooking fuel use, proximity to environmental hazards, occupational exposures and dietary patterns; cardiovascular and metabolic measurements, including blood pressure, body mass index, blood glucose and serum lipid profiles; serological data; interview metadata, including interviewer identifier, date of administration and language; and a biospecimen registry recording tube type, volume, storage location, aliquot number and freeze-thaw cycles for all archived samples. Across all study arms, data structures are harmonised using a shared participant identifier system, enabling linkage of records for individuals represented in both the community and hospital datasets.

### 6.2 Planned Analyses

The SAHEL Study is designed to support a phased analytical program spanning epidemiology, biomarker science, genomics, environmental health and longitudinal follow-up. An overview of this phased program is shown in Figure 8.

**Figure 8.**
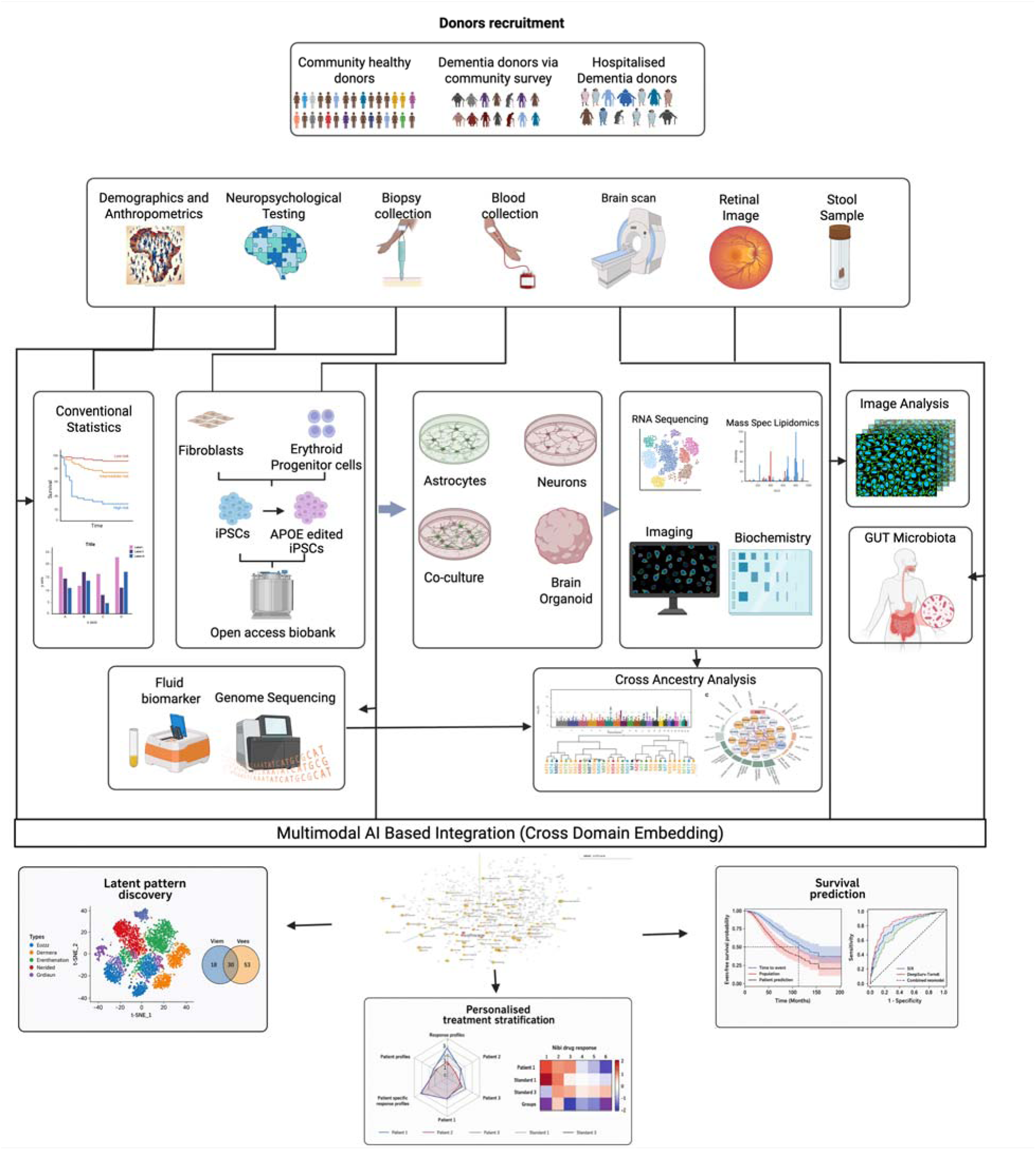
Planned analytical program for the SAHEL Study. Ten data-generating and analytical domains; epidemiological characterization, biomarker profiling, genomic and ancestry analyses, iPSC molecular analyses, environmental genomics, molecular modeling, longitudinal follow-up, gut microbiome profiling, neuroimaging, and ophthalmic and retinal analyses, feed into a multimodal AI-based integration framework to support latent pattern discovery, prediction of longitudinal outcomes, and identification of clinically relevant participant subgroups.

Phase 1 (Epidemiological characterization) will characterize the burden, distribution and determinants of cognitive impairment in north-eastern Nigeria, including analyses of environmental and vascular risk factors and stratification by major ethnolinguistic groups where sample sizes permit.

Phase 2 (Biomarker profiling) will include lipidomic, metabolomic and proteomic profiling of archived serum and plasma samples, alongside inflammatory marker panels and serological testing for common chronic viral infections. Candidate blood-based biomarkers, including the amyloid-beta (Aβ)42/40 ratio, phosphorylated tau species such as p-tau181 and p-tau217, neurofilament light chain (NfL), microtubule-binding region tau (MTBR-tau) and glial fibrillary acidic protein (GFAP), will be quantified using ultrasensitive proteomic approaches in collaboration with Dr Thomas Karikari at the University of Pittsburgh, enabling systematic evaluation of these markers in an African-ancestry neurodegeneration cohort.

Phase 3 (Genomic and ancestry analyses) will initially employ genome-wide genotyping arrays to evaluate established AD-associated loci, including APOE, BIN1, CLU and PICALM, alongside genome-wide analyses of population structure and admixture across major ethnolinguistic groups, including Kanuri, Fulani and Hausa participants. Whole-genome sequencing of community and hospital participants is planned as a future phase, pending dedicated sequencing funding, and will enable more comprehensive discovery-level analyses of genetic variation in these underrepresented populations.

Phase 4 (iPSC molecular analyses) will use iPSC-derived neurons, astrocytes and cerebral organoids from SAHEL donors to investigate how genetic and ancestral variation influences cellular and molecular pathways relevant to AD and related dementias, including amyloid-beta processing, tau pathology and cellular lipid homeostasis, using RNA sequencing, proteomics and lipidomics. Initial studies will include investigation of APOE-associated biology while enabling analysis of additional genetic risk and protective factors identified through genomic studies.

Phase 5 (Environmental genomics) will integrate environmental exposure data with genomic and epigenomic data, including analyses of gene–environment interactions involving potential neurotoxicant exposures and epigenome-wide association studies (EWAS) of environmental exposures and cognitive outcomes.

Phase 6 (Molecular modeling) will encompass computational modeling of disease-related networks integrating genomic variation, environmental exposures and clinical phenotypes, alongside causal inference approaches, including Mendelian randomisation where suitable genetic instruments are available, to investigate potentially causal relationships involving environmental and metabolic risk factors.

Phase 7 (Gut microbiota) will characterize gut microbiota composition and diversity from stool specimens collected during longitudinal follow-up and investigate associations with environmental exposures, dietary patterns, genetic variation and cognitive status.

Phase 8 (Neuroimaging analyses) will use structural MRI and, where available, diffusion and functional MRI to quantify markers of neurodegeneration and cerebrovascular disease, including regional brain volumes, cortical thickness and white matter hyperintensity burden, enabling systematic neuroimaging characterization of dementia and cognitive impairment in this population.

Phase 9 (Ophthalmic and retinal analyses) will use retinal imaging acquired with the Topcon Maestro2 platform, including macular and wide-field optical coherence tomography (OCT), OCT angiography and color fundus photography, to characterize retinal microvascular and structural features associated with cognitive impairment, dementia and systemic vascular disease, and to investigate genetic susceptibility to age-related retinal and ocular disorders, including age-related macular degeneration and glaucoma.

Phase 10 (Multimodal AI-based integration) will apply multimodal artificial intelligence approaches across cohort data domains, including genomic, environmental, clinical, biomarker, neuroimaging, retinal and cellular/omics data, to identify multidimensional patterns and relationships not readily captured using conventional statistical approaches and to develop and evaluate models for disease classification, risk stratification and prediction of longitudinal cognitive and clinical outcomes.

### 6.3 Data Access and Collaboration

The SAHEL Study is committed to open science, equitable collaboration and responsible data sharing. We welcome enquiries from researchers, clinicians and public health practitioners interested in collaborating on analyses of SAHEL data or biospecimens. Collaboration proposals will be reviewed by the SAHEL Steering Committee, with priority given to projects that (1) advance scientific understanding of dementia and related conditions in African populations, (2) involve Nigerian researchers in substantive scientific roles, and (3) contribute to research capacity strengthening in north-eastern Nigeria.

Data access requests should be directed to the corresponding author. De-identified analytical datasets underlying published analyses will be made available through appropriate controlled-access mechanisms, where permitted by participant consent and applicable ethical and regulatory requirements, with access governed by SAHEL data-access procedures and in accordance with FAIR data principles. Access to biospecimens for external collaborative projects will follow a separate protocol overseen by the SAHEL Biobank Committee. External requests for data or biospecimens will be subject to appropriate scientific and governance review and, where required, approval by relevant research ethics committees. These procedures are intended to protect participant confidentiality, ensure responsible scientific use and maintain appropriate stewardship of a resource developed in partnership with communities in north-eastern Nigeria.

## 7. Discussion

### 7.1 Principal Findings and Scientific Significance

The SAHEL Study has established a deeply phenotyped, prospective, multi-arm aging and dementia cohort in north-eastern Nigeria, enrolling more than 1,000 community-dwelling older adults and over 100 hospital-based participants with cognitive impairment or suspected dementia from several ethnolinguistically diverse populations.

The first principal scientific asset of SAHEL is its ancestral uniqueness. The Kanuri, Fulani, Hausa, Shuwa Arab, and Babur/Bura peoples of the Lake Chad Basin region represent populations with diverse ancestral histories, migration patterns, cultural diversities and civilisational exchange. The Kanuri, in particular, descendants of the Kanem-Bornu Empire, one of the longest-lasting states in the Central Sudan in African history, are substantially underrepresented in existing dementia genomics datasets. SAHEL provides the first opportunity to investigate how ancestry-related factors influence neurodegenerative disease risk and resilience in these populations. More broadly, the sociocultural and genomic diversity of the Lake Chad Basin region offers a distinctive opportunity to examine how ancestry-related genomic factors interact with environment and culture to shape neurodegenerative disease risk and resilience.[41]

The second asset is the detailed characterization of potential environmental exposures. SAHEL participants experience several potential routes of exposure relevant to neurological health, including reliance on groundwater, household air pollution associated with biomass fuel use, proximity to agrochemical activities and reported occupational toxicant exposures. These exposures have plausible or established links to neurological and systemic health outcomes but remain comparatively understudied in dementia cohorts in sub-Saharan Africa. The environmental characterization embedded within SAHEL provides an opportunity to investigate whether measured environmental exposures are associated with cognitive impairment, biomarkers and neurodegenerative outcomes [42]

The third asset is the substantial burden of positive cognitive screening outcomes identified at baseline. Mini-Cog positivity was observed in 36.3% of the community cohort. However, because Mini-Cog performance may be influenced by educational and sociocultural factors, particularly in populations with limited formal Western education, this figure should not be interpreted as the prevalence of clinical cognitive impairment or dementia. Ongoing detailed cognitive and clinical assessments will establish the extent to which screen positivity corresponds to clinically defined cognitive impairment. If a high burden is confirmed, this would have important implications for dementia detection, health-service planning and community education in north-eastern Nigeria.

The fourth asset is the SAHEL biobank, comprising cryopreserved serum, plasma and buffy coat from more than 900 participants across the community and hospital arms. This constitutes an important biospecimen resource for biomarker, genomic and other molecular studies and enables future evaluation of emerging dementia biomarkers as analytical technologies advance. Given the substantial underrepresentation of African populations in dementia biomarker development and validation studies, SAHEL provides an opportunity to improve the representation of African populations in biomarker research.

The fifth asset is the African Somatic and Stem Cell Bank, through which iPSC lines generated from African donors are being developed as an openly accessible resource distributed across collaborating repositories in Nigeria, the United Kingdom and the United States. These cellular models provide a platform for investigating how genetic and ancestral variation influences molecular and cellular pathways relevant to Alzheimer’s disease and related dementias. Initial studies will investigate APOE-associated biology, while the resource will enable future investigation of additional genetic risk and protective factors.

The sixth, planned asset is the SAHEL gut microbiota sub-study. Stool specimens, to be collected prospectively at longitudinal follow-up, will generate the first reference gut microbiota data for the Kanuri, Fulani, Hausa, Babur/Bura, and Shuwa Arab peoples, and test the relationships among the gut microbiota, diet, environmental exposures, genetic variation and cognitive outcomes in this population [18].

The Seventh planned asset is deep ophthalmic phenotyping: comprehensive retinal imaging will enable investigation of retinal microvascular and neurodegenerative markers associated with dementia and systemic vascular disease, while also supporting studies of age-related macular degeneration and glaucoma susceptibility loci in a population where such data are virtually absent and disease prevalence patterns differ markedly from European-ancestry cohorts.

### 7.2 Context in the Global Dementia Research Landscape

SAHEL joins a small but growing number of African aging and dementia research cohorts and initiatives, including the EPOCH Study in Ghana, INDEPTH Network studies across Ghana, South Africa and Kenya, the Africa Wits-INDEPTH Partnership for Genomic Studies (AWI-Gen), the African Dementia Consortium (AfDC), and ongoing cohorts in Kenya harmonised through the Africa-FINGERS network’s Kenya site at Aga Khan University, Nairobi (Section 4.2). Unlike many existing African dementia studies, which have been concentrated in particular regions and populations, SAHEL captures ethnolinguistically and ancestrally diverse populations from the Lake Chad Basin, including Kanuri, Fulani, Hausa, Shuwa Arab and Babur/Bura peoples, who remain substantially underrepresented in global dementia research. Several additional features distinguish SAHEL from existing African aging studies: its geographic focus on north-eastern Nigeria, a region with very limited representation in dementia research; the ancestral and sociocultural diversity of its sample; its integration of environmental, metabolic, genomic and clinical phenotyping within a single cohort framework; its two-arm design enabling integration of community- and hospital-based data; and the establishment of structured biospecimen and iPSC biobanking platforms within a resource-limited setting.

### 7.3 Limitations

The SAHEL Study has several important limitations. First, the cross-sectional nature of the current data precludes assessment of incidence, temporal relationships, and within-person cognitive change; follow-up at 12-month intervals is underway and will provide longitudinal observations. Second, performance on cognitive screening instruments may have been influenced by low levels of formal educational attainment and cultural differences in test familiarity.

Follow-on clinical assessments in the community arm will provide more robust estimates of the frequency of cognitive impairment and dementia, as well as an opportunity to examine the performance of the Mini-Cog in this setting. Biospecimen collection, while achieving high coverage (77.5% for venous blood in the community arm), was incomplete for a proportion of participants, and the determinants of non-collection warrant analysis to assess potential selection bias. Hausa is widely spoken across the region as a lingua franca, and validated Hausa-language versions of the clinical assessment instruments used in this study were available, which is expected to mitigate some linguistic and cultural validity concerns among Hausa-speaking participants. However, formal cultural validation data for the Kanuri and Fulfulde language adaptations of several instruments remain limited, and future validation work is needed. Finally, specialist clinical assessment was completed in only 21 of 102 enrolled hospital participants at the time of analysis, limiting the diagnostic characterization of the hospital cohort; estimates reported from this cohort should therefore be considered preliminary.

### 7.4 Capacity Building and Global Health Impact

A defining component of the SAHEL Study is its commitment to building scientific capacity in north-eastern Nigeria. The study has trained a cadre of field researchers, nurses, phlebotomists, and data managers in research methods, cognitive assessment, biospecimen handling, data collection, and the ethical conduct of research involving human participants. The establishment of functional biobanking infrastructure in the region represents a durable research asset that will extend beyond the current study phase and enable future research activities. These capacity investments are designed to create a sustainable research infrastructure capable of supporting subsequent cohort phases, collaborative studies, and the training of the next generation of Nigerian dementia researchers, independently of any single funding cycle.

## 8. Conclusions

The SAHEL Study has established a deeply phenotyped, prospective, multi-arm aging and dementia cohort in north-eastern Nigeria, incorporating populations from the Lake Chad Basin that remain markedly underrepresented in dementia research. Its integration of systematic environmental assessment, multidomain clinical phenotyping, biospecimen banking, genomic analyses, and an open-access iPSC resource provides a platform for investigating dementia and its determinants in this understudied region. The high proportion of community participants screening positive for cognitive concern (36.3%), alongside substantial reported environmental exposures of potential neurotoxicological relevance, highlights the importance of longitudinal investigation to determine their clinical significance and relationship to subsequent cognitive outcomes. Continued follow-up and collaborative analyses integrating clinical, biomarker, genomic, environmental, neuroimaging, and iPSC data will enable estimation of dementia incidence and investigation of risk factors and gene–environment interactions among Kanuri, Fulani, Hausa, and other populations represented in the cohort. In doing so, SAHEL has the potential to address an important gap in the global evidence base on dementia and contribute evidence relevant to dementia prevention, diagnosis, and care in north-eastern Nigeria and other African settings.

## 9. Collaborating Institutions and Investigators

The SAHEL Study brings together a collaborative research network spanning Nigeria, Ghana, The Gambia, the United Kingdom, the United States, Canada, and the Netherlands. The network was assembled to integrate expertise in clinical phenotyping, dementia epidemiology, fluid biomarkers, neuroimaging, genomics, iPSC generation and characterization, lipidomics, bioinformatics, and computational analysis, while maintaining Nigerian scientific leadership across the research program.

### 9.1 Nigerian Clinical and Research Sites

Clinical recruitment and research activities are coordinated through the Northern Nigeria Dementia Research Group (NNDRG), a collaborative network established by the principal investigator across six participating institutions and sites in north-eastern Nigeria: the Federal Neuropsychiatric Hospital Maiduguri (FNPHM), the primary hospital-arm recruitment site and a major neuropsychiatric referral center in the region; Yobe State University Teaching Hospital (YSUTH), Damaturu, which serves as a clinical recruitment site and the primary neuroimaging site for the cohort; Yobe State Specialist Hospital Damaturu; Yobe State Specialist Hospital Potiskum; Yobe State Specialist Hospital Gashua; and the Biomedical Science Research and Training Centre (BioRTC) at Yobe State University. The three Specialist Hospitals support community and clinical recruitment across different parts of Yobe State, extending the geographical reach of the cohort. BioRTC, established at Yobe State University with support from the Yobe State Government, serves as the central research laboratory and primary Nigerian biobanking site, supporting biospecimen processing and storage, fibroblast culture, iPSC generation and characterization, and cell-based studies.

### 9.2 International Collaborating Investigators

The SAHEL Study is supported by international collaborators contributing complementary expertise across its major scientific components. Dr Thomas Karikari (University of Pittsburgh, USA) leads plasma biomarker analyses using ultrasensitive biomarker platforms. Prof. Celeste Karch (Washington University in St. Louis, USA) supports iPSC generation, characterization, biobanking, and downstream cellular studies. Dr Chiadi Onyike (Johns Hopkins University, USA) contributes expertise in dementia epidemiology, community screening, clinical assessment, and training. Dr Udunna Anazodo (McGill University, Canada) leads the neuroimaging component of the study. Dr Rik van der Kant (Vrije Universiteit Amsterdam, Netherlands) supports lipidomics analyses, while Dr Abdul Karim Sesay (MRC Unit The Gambia) and Dr Yaw Aniweh (WACCBIP, University of Ghana) contribute expertise in genomics and bioinformatics. The program is further supported by an independent Scientific Advisory Board and a Data Governance and Management Committee, whose members provide complementary expertise in Alzheimer’s disease and dementia research, epidemiology, statistics, genomics, bioethics, data governance, and community-based research.

### 9.3 Biobank Network

The SAHEL biobank uses a distributed model designed to maintain biospecimen resources within Nigeria while enabling specialised collaborative analyses. BioRTC serves as the primary biobank and laboratory site, maintaining cryopreserved blood-derived biospecimens, fibroblasts, erythroid progenitor cells, and iPSC lines. Sussex Neuroscience at the University of Sussex serves as a UK collaborative site for iPSC lines and derivative cellular models, while Washington University in St. Louis serves as a US collaborative site supporting iPSC-related studies and associated molecular analyses. Biospecimen sharing between participating institutions is governed by appropriate Material Transfer Agreements and study governance procedures, with BioRTC maintaining oversight of the primary resource. These arrangements are designed to ensure participant confidentiality, ethical compliance, appropriate use of samples, and traceability of materials shared across collaborating institutions.

### 9.4 Assessment, Data, and Biospecimen Pipelines

SAHEL uses defined assessment, data, and biospecimen pipelines linking participant recruitment and field collection to central processing and specialised analyses. Cognitive, functional, demographic, environmental, and clinical data are captured electronically at the point of contact using KoBoCollect, with QField additionally used for household mapping. Data are transmitted via encrypted connections to the secure central database described in Section 6.1 and subsequently harmonised into study-wide analytical datasets. Access to study data by collaborating investigators is subject to the study’s governance and data-access procedures.

Biospecimens follow parallel workflows from participant collection through local processing, biobanking, and specialised analysis. Venous blood is processed and aliquoted locally, with designated plasma and serum aliquots available for ultrasensitive biomarker analyses in Dr Thomas Karikari’s laboratory at the University of Pittsburgh. Skin biopsies are cultured locally, with fibroblasts reprogrammed into iPSCs at BioRTC; selected lines and derivative cellular models are subsequently shared with collaborating laboratories, including the University of Sussex and Washington University in St. Louis, for specialised cellular and molecular studies. DNA derived from blood samples supports planned genomic analyses in collaboration with Dr Yaw Aniweh, Dr Abdul Karim Sesay and other genomic partners.

All biospecimen transfers are recorded within the study’s biospecimen management system and linked to the originating participant through coded study identifiers. Transfers to collaborating institutions are conducted under applicable Material Transfer Agreements and study governance approvals, providing traceability while protecting participant confidentiality.

## Data Availability

The data generated and analysed in this study are not publicly available because they contain sensitive participant-level information and are subject to ethical, consent and data-governance requirements. De-identified data may be made available to qualified researchers following a reasonable request and review and approval by the appropriate SAHEL Study governance committee, in accordance with participant consent, applicable ethical approvals and Nigerian data-governance requirements.

## Acknowledgements

We are gratefully to all participants and their families for their time, trust and contribution to the SAHEL Study. We thank the community and religious leaders, Primary Health Care staff, field researchers, community mobilisers, nurses, phlebotomists, laboratory personnel, data managers and clinical teams whose contributions made community and hospital recruitment possible. We particularly acknowledge the support of the staff from Federal Neuropsychiatric Hospital Maiduguri, Yobe State University Teaching Hospital, Yobe State Specialist Hospitals, and the Biomedical Science Research and Training Centre (BioRTC), in supporting participant recruitment, clinical assessment, biospecimen collection, processing and biobanking. The study benefited substantially from engagement with local communities in Yobe and Borno States, whose partnership has been integral to the development of the SAHEL research programme. We also acknowledge the wider Northern Nigeria Dementia Research Group and the scientific, clinical and technical collaborators contributing to the continued development of the SAHEL platform.

## Funding

This work was principally supported by a Wellcome Trust Career Development Award to Mahmoud Bukar Maina (224493/Z/21/Z). Earlier support from the Rainwater Charitable Foundation and the Alzheimer’s Association (AARFD-22-923450) contributed to the establishment of the Northern Nigerian Dementia Research Group and the foundational development of the SAHEL cohort. We also acknowledge the support of the Yobe State Government for the establishment and continued development of BioRTC and the research infrastructure supporting the SAHEL Study.

### Conflict of Interest

The authors declare no conflicts of interest.

### Author contributions

Conceptualisation: MBM, CUO. Methodology: MBM, SHK, IAW, BWG, UBM, PNO, MYM, MAF, MMG, LB, ZM, BKM, CSL, OO, YA, TY, UA, CUM, AUF, HJI. Investigation: MBM, SHK, HJI, AUF, IAW, UBM, PNO, MYM, MAF, MMG, MMA, ZUA, MKS, ZBY, FMK, NMS, PD, LEN, LB, ZM, DU, AIA, SJ, OO. Clinical assessment and phenotyping: IAW, UBM, PNO, MYM, MAF, MMA, ZUA, MKS, ZBY, FMK, NMS, PD, LEN, SJ. Data curation and management: MBM, SHK, MMG, LB, MMA, ZUA, MKS, ZBY, FMK, NMS, PD, LEN, SJ. Formal analysis: MBM, SHK. Resources: MBM, IAW, UBM, PNO, BWG, BKM, CSL, YA, RLG, TY, UA, CUM, TKK, CMK, CUO. Supervision: CUO, MBM. Project administration: MBM, SHK, IAW, CUO, MYM. Funding acquisition: MBM. Writing - original draft: SHK, MBM. Writing – review and editing: All authors. All authors read and approved the final manuscript.

## Abbreviations

AD: Alzheimer’s disease
BADL: Basic Activities of Daily Living
BMI: body mass index
CDT: Clock Drawing Test
EWAS: Epigenome-Wide Association Study
FNPHM: Federal Neuropsychiatric Hospital Maiduguri
GFAP: Glial Fibrillary Acidic Protein
GWAS: Genome-Wide Association Study
IADL: Instrumental Activities of Daily Living
LGA: Local Government Area
MCI: Mild Cognitive Impairment
Mini-Cog: Mini-Cognitive Assessment Instrument
MoCA: Montreal Cognitive Assessment
NfL: Neurofilament Light Chain
NPI-Q: Neuropsychiatric Inventory Questionnaire
PHQ-2: Patient Health Questionnaire (2-item)
QDRS: Quick Dementia Rating System
SAHEL: Study of Ancestry, Health, Environment, and Late-Life Neurodegeneration
SST: Serum Separator Tube
VaD: Vascular Dementia
ZBI: Zarit Burden Interview.

## Notes

### Competing Interest Statement

The authors have declared no competing interest.

### Author Declarations

The Institutional Research and Ethical Board of the Federal Neuropsychiatric Hospital Maiduguri gave ethical approval for the hospital-based component of this work (FNPH/082023/REC140). The State Health Research Ethics Committee of the Yobe State Ministry of Health gave ethical approval for the community-based component of this work (MOH/GEN/747/Vol. 1).

## References

[1] Organization WH. Global status report on the public health response to dementia 2021.

[2] Nichols E, Steinmetz JD, Vollset SE, Fukutaki K, Chalek J, Abd-Allah F, et al. Estimation of the global prevalence of dementia in 2019 and forecasted prevalence in 2050: an analysis for the Global Burden of Disease Study 2019. Lancet Public Health 2022;7:e105– 25.

[3] Ferri CP, Prince M, Brayne C, Brodaty H, Fratiglioni L, Ganguli M, et al. Global prevalence of dementia: a Delphi consensus study. The Lancet 2005;366:2112–7.

[4] Fatumo S, Chikowore T, Choudhury A, Ayub M, Martin AR, Kuchenbaecker K. A roadmap to increase diversity in genomic studies. Nat Med 2022;28:243–50.

[5] Joshi E, Biddanda A, Popoola J, Yakubu A, Osakwe O, Attipoe D, et al. Whole-genome sequencing across 449 samples spanning 47 ethnolinguistic groups provides insights into genetic diversity in Nigeria. Cell Genomics 2023;3.

[6] Fortes-Lima CA, Diallo MY, Janoušek V, Černý V, Schlebusch CM. Population history and admixture of the Fulani people from the Sahel. The American Journal of Human Genetics 2025;112:261–75.

[7] Černý V, Salas A, Hájek M, Žaloudková M, Brdička R. A bidirectional corridor in the Sahel Sudan belt and the distinctive features of the Chad Basin populations: a history revealed by the mitochondrial DNA genome. Ann Hum Genet 2007;71:433–52.

[8] Duncan L, Shen H, Gelaye B, Meijsen J, Ressler K, Feldman M, et al. Analysis of polygenic risk score usage and performance in diverse human populations. Nat Commun 2019;10:3328.

[9] Gauthier S, Rosa-Neto P, Morais JA, Webster C. World Alzheimer Report 2021: Journey through the diagnosis of dementia. Alzheimer’s Disease International 2021;2022:30.

[10] Livingston G, Huntley J, Liu KY, Costafreda SG, Selbæk G, Alladi S, et al. Dementia prevention, intervention, and care: 2024 report of the Lancet standing Commission. The Lancet 2024;404:572–628.

[11] Hendrie HC, Ogunniyi A, Hall KS, Baiyewu O, Unverzagt FW, Gureje O, et al. Incidence of dementia and Alzheimer disease in 2 communities: Yoruba residing in Ibadan, Nigeria, and African Americans residing in Indianapolis, Indiana. JAMA 2001;285:739–47.

[12] Gómez-Olivé FX, Montana L, Wagner RG, Kabudula CW, Rohr JK, Kahn K, et al. Cohort profile: health and ageing in Africa: a longitudinal study of an INDEPTH community in South Africa (HAALSI). Int J Epidemiol 2018;47:689–690j.

[13] Prince M, Ferri CP, Acosta D, Albanese E, Arizaga R, Dewey M, et al. The protocols for the 10/66 dementia research group population-based research programme. BMC Public Health 2007;7:165.

[14] Feil R, Fraga MF. Epigenetics and the environment: emerging patterns and implications. Nature Reviews Genetics 2012 13:2 2012;13:97–109. 10.1038/nrg3142.

[15] Ahmed SD, Agodzo SK, Adjei KA, Deinmodei M, Ameso VC. Preliminary investigation of flooding problems and the occurrence of kidney disease around Hadejia-Nguru wetlands, Nigeria and the need for an ecohydrology solution. Ecohydrology & Hydrobiology 2018;18:212–24. 10.1016/J.ECOHYD.2017.11.005.

[16] Aborode AT, Otorkpa OJ, Abdullateef AO, Oluwaseun OS, Adegoye GA, Aondongu NJ, et al. Impact of Climate Change-Induced Flooding Water Related Diseases and Malnutrition in Borno State, Nigeria: A Public Health Crisis. Environ Health Insights 2025;19:11786302251321684. 10.1177/11786302251321683.

[17] Wakawa IA, Musami UB, Kwairanga SH, Ogualili PN, Mahmood MY, Fugu MA, et al. Dementia in a resource constrained sub Saharan African setting: A comprehensive retrospective analysis of prevalence, risk factors, and management at the only neuropsychiatric facility in Northeastern Nigeria. Alzheimer’s & Dementia 2025;21. 10.1002/alz.14538.

[18] Seo DO, O’Donnell D, Jain N, Ulrich JD, Herz J, Li Y, et al. ApoE isoform- and microbiota-dependent progression of neurodegeneration in a mouse model of tauopathy. Science (1979) 2023;379. 10.1126/SCIENCE.ADD1236;PAGE:STRING:ARTICLE/CHAPTER.

[19] Mohammed Said J, Jibril A, Isah R, Beida O. Pattern of Presentation and Utilization of Services for Mental and Neurological Disorders in Northeastern Nigeria: A Ten Year Study. Psychiatry J 2015;2015:328432.

[20] Borson S, Scanlan J, Brush M, Vitaliano P, Dokmak A. The mini cog: a cognitive ‘vital signs’ measure for dementia screening in multi lingual elderly. Int J Geriatr Psychiatry 2000;15:1021–7.

[21] Riley McCarten J, Anderson P, Kuskowski MA, McPherson SE, Borson S, Dysken MW. Finding dementia in primary care: the results of a clinical demonstration project. J Am Geriatr Soc 2012;60:210–7.

[22] Udeh-Momoh CT, Maina R, Anazodo UC, Akinyemi R, Atwoli L, Baker L, et al. Dementia risk reduction in the African context: Multi-national implementation of multimodal strategies to promote healthy brain aging in Africa (the Africa-FINGERS project). Alzheimer’s & Dementia 2024;20:8987–9003. 10.1002/ALZ.14344.

[23] Baiyewu O, Unverzagt FW, Lane KA, Gureje O, Ogunniyi A, Musick B, et al. The Stick Design test: A new measure of visuoconstructional ability. Journal of the International Neuropsychological Society 2005;11:598–605. 10.1017/S135561770505071X.

[24] Paddick SM, Gray WK, Ogunjimi L, Lwezuala B, Olakehinde O, Kisoli A, et al. Validation of the Identification and Intervention for Dementia in Elderly Africans (IDEA) cognitive screen in Nigeria and Tanzania. BMC Geriatrics 2015 15:1 2015;15:53-. 10.1186/S12877-015-0040-1.

[25] Julayanont P, Phillips N, Chertkow H, Nasreddine ZS. Montreal Cognitive Assessment (MoCA): Concept and Clinical Review. Cognitive Screening Instruments 2013:111–51. 10.1007/978-1-4471-2452-8_6.

[26] Nasreddine ZS, Phillips NA, Bedirian V, Charbonneau S, Whitehead V, Collin I, et al. The Montreal Cognitive Assessment, MoCA: A Brief Screening Tool For Mild Cognitive Impairment. J Am Geriatr Soc 2005;53:695–9.

[27] Galvin JE. The Quick Dementia Rating System (QDRS): A rapid dementia stagingtool. Alzheimer’s and Dementia: Diagnosis, Assessment and Disease Monitoring 2015;1:249–59. 10.1016/J.DADM.2015.03.003;WGROUP:STRING:PUBLICATION.

[28] Kaufer DI, Cummings JL, Ketchel P, Smith V, MacMillan A, Shelley T, et al. Validation of the NPI-Q, a brief clinical form of the Neuropsychiatric Inventory. J Neuropsychiatry Clin Neurosci 2000;12:233–9. 10.1176/JNP.12.2.233.

[29] Wei L, Hodgson C. Clinimetrics: The Lawton-Brody Instrumental Activities of Daily Living Scale. J Physiother 2023;69:57. 10.1016/J.JPHYS.2022.06.007.

[30] Gratão ACM, Brigola AG, Ottaviani AC, Luchesi BM, Souza ÉN, Rossetti ES, et al. Brief version of Zarit Burden Interview (ZBI) for burden assessment in older caregivers. Dement Neuropsychol 2019;13:122–9. 10.1590/1980-57642018DN13-010015.

[31] Muhammad Z, Brown PW, Babazau L, Alkhamis AI, Goni BW, Nggada HA, et al. Generation of an induced pluripotent stem cell line from a healthy adult indigenous Nigerian participant 2023. 10.1101/2023.07.21.550059.

[32] Beam CR, Kaneshiro C, Jang JY, Reynolds CA, Pedersen NL, Gatz M. Differences between women and men in incidence rates of dementia and Alzheimer’s disease. Journal of Alzheimer’s Disease 2018;64:1077–83.

[33] Organization WH. The global dementia observatory reference guide. World Health Organization; 2018.

[34] Paddick S-M, Gray WK, Ogunjimi L, Lwezuala B, Olakehinde O, Kisoli A, et al. Validation of the Identification and Intervention for Dementia in Elderly Africans (IDEA) cognitive screen in Nigeria and Tanzania. BMC Geriatr 2015;15:53.

[35] Perlman CM, Hirdes JP, Barbaree H, Fries BE, McKillop I, Morris JN, et al. Development of mental health quality indicators (MHQIs) for inpatient psychiatry based on the interRAI mental health assessment. BMC Health Serv Res 2013;13:15.

[36] Brown C, Barner J, Bohman T, Richards K. A multivariate test of an expanded Andersen Health Care utilization model for complementary and alternative medicine (CAM) use in African Americans. The Journal of Alternative and Complementary Medicine: Paradigm, Practice, and Policy Advancing Integrative Health 2009;15:911–9.

[37] Giwa AS, Memon AG, Ahmad J, Ismail T, Abbasi SA, Kamran K, et al. Assessment of high fluoride in water sources and endemic fluorosis in the North-Eastern communities of Gombe State, Nigeria. Environmental Pollutants and Bioavailability 2021;33:31–40. 10.1080/26395940.2021.1908849.

[38] Emenike PGC, Tenebe I, Ogarekpe N, Omole D, Nnaji C. Probabilistic risk assessment and spatial distribution of potentially toxic elements in groundwater sources in Southwestern Nigeria. Scientific Reports 2019 9:1 2019;9:15920-. 10.1038/s41598-019-52325-z.

[39] Oudin A, Segersson D, Adolfsson R, Forsberg B. Association between air pollution from residential wood burning and dementia incidence in a longitudinal study in Northern Sweden. PLoS One 2018;13:e0198283. 10.1371/JOURNAL.PONE.0198283.

[40] Onakpa MM, Njan AA, Kalu OC. A Review of Heavy Metal Contamination of Food Crops in Nigeria. Ann Glob Health 2018;84:488–94. 10.29024/AOGH.2314.

[41] Ceesay H. A History of Borno: Trans-Saharan African Empire to Failing Nigerian State by Vincent Hiribarren (review). Journal of Global South Studies 2019;36:216–7. 10.1353/GSS.2019.0013.

[42] Antoniadou F, Papamitsou T, Kavvadas D, Kapoukranidou D, Sioga A, Papaliagkas V. Toxic Environmental Factors and their Association with the Development of Dementia: a Mini Review on Heavy Metals and Ambient Particulate Matter. Mater Sociomed 2020;32:299. 10.5455/MSM.2020.32.299-306.

[43] Maina MB, et al. Somatic and Stem Cell Bank to study the contribution of African ancestry to dementia: African iPSC Initiative. Alzheimer’s & Dementia. 2025;21:e70145. doi:10.1002/alz.70145.

[44] Zhou B, Perel P, Mensah GA, Ezzati M, et al. Global epidemiology, health burden and effective interventions for elevated blood pressure and hypertension. Nature Reviews Cardiology. 2021;18:785–802. doi:10.1038/s41569-021-00559-8.

